# Adaptive metrics for an evolving pandemic A dynamic approach to area-level COVID-19 risk designations

**DOI:** 10.1101/2023.02.15.23285969

**Authors:** Alyssa M. Bilinski, Joshua A. Salomon, Laura A. Hatfield

**Affiliations:** Departments of Health Services, Policy and Practice & Biostatistics, Brown University, 121 S. Main St., Providence, RI 02912 USA; Department of Health Policy, Stanford University, Stanford, CA 94305 USA; Department of Health Care Policy, Harvard Medical School, 180 Longwood Ave., Boston, MA 02115 USA

**Author notes:** A.M.B designed research and analyzed data; A.M.B., J.A.S., and L.A.H. interpreted results and wrote the paper. The authors declare no competing interests.

**Keywords:** Infectious disease dynamics, Decision theory, Risk prediction, COVID-19

## Abstract

Throughout the COVID-19 pandemic, policymakers have proposed risk metrics, such as the CDC Community Levels, to guide local and state decision-making. However, risk metrics have not reliably predicted key outcomes and often lack transparency in terms of prioritization of false positive versus false negative signals. They have also struggled to maintain relevance over time due to slow and infrequent updates addressing new variants and shifts in vaccine- and infection-induced immunity. We make two contributions to address these weaknesses of risk metrics. We first present a framework to evaluate predictive accuracy based on policy targets related to severe disease and mortality, allowing for explicit preferences toward false negative versus false positive signals. This approach allows policymakers to optimize metrics for specific preferences and interventions. Second, we propose a novel method to update risk thresholds in real-time. We show that this adaptive approach to designating areas as “high risk” improves performance over static metrics in predicting 3-week-ahead mortality and intensive care usage at both state and county levels. We also demonstrate that with our approach, using only new hospital admissions to predict 3-week-ahead mortality and intensive care usage has performed consistently as well as metrics that also include cases and inpatient bed usage. Our results highlight that a key challenge for COVID-19 risk prediction is the changing relationship between indicators and outcomes of policy interest. Adaptive metrics therefore have a unique advantage in a rapidly evolving pandemic context.

**Significance Statement:** In the rapidly-evolving COVID-19 pandemic, public health risk metrics often become less relevant over time. Risk metrics are designed to predict future severe disease and mortality based on currently-available surveillance data, such as cases and hospitalizations. However, the relationship between cases, hospitalizations, and mortality has varied considerably over the course of the pandemic, in the context of new variants and shifts in vaccine- and infection-induced immunity. We propose an adaptive approach that regularly updates metrics based on the relationship between surveillance inputs and future outcomes of policy interest. Our method captures changing pandemic dynamics, requires only hospitalization input data, and outperforms static risk metrics in predicting high-risk states and counties.

---

**U**nderstanding the evolution of infectious disease risk is critical for individuals making decisions about personal precautions, policymakers recommending mitigation measures, and health care institutions planning for future surges. Throughout the COVID-19 pandemic, indicators such as reported cases and percent of PCR tests positive for SARSCoV-2 have been used to guide pandemic response (1–4). Currently, the Center for Disease Control and Prevention (CDC)’s Community Levels designate areas as low, medium, or high risk based on reported cases, new COVID-19 hospital admissions, and percentage of inpatient beds occupied by COVID-19 patients (2).

However, COVID-19 risk metrics have had several weaknesses. First, policymakers have struggled to identify leading indicators of key health outcomes. For example, PCR test positivity was abandoned as a trigger for school closures because it did not reliably predict in-school transmission (5). Community metrics have focused on predicting severe disease and mortality (2, 6). For example, the indicators used in CDC Community Levels were selected because they correlated with ICU rates and mortality 3 weeks in the future (2). However, the thresholds for low, medium, and high did not correspond to specific future mortality rates (7), thus complicating the understanding of a “high risk” designation.

Second, many metrics fail to distinguish different error types. Falsely classifying an area as high risk may prompt unnecessary or harmful interventions, while a false negative may fail to activate needed public health measures (8). Individuals and policymakers may vary in their preferences for avoiding these two types of errors, but current methods fail even to make these preferences explicit (9).

Finally, changes in available data, COVID-19 variants, and levels of immunity can render metrics obsolete as the pandemic evolves (10). For instance, with the omicron variant, cases and hospital admissions have corresponded to lower levels of mortality than in earlier waves. Shifts from PCR to at-home testing and changes in case reporting have also made case data less reliable and available over time (11, 12). Ad hoc updates to risk designations are insufficient to ensure that the metrics remain relevant. Moreover, transparency in the process is key to alleviating concerns about “moving the goalposts” (13).

This paper makes two contributions to address these weaknesses in the context of COVID-19 community risk metrics. First, we propose a framework for predictive accuracy that incorporates preferences over false negatives versus false positives, using weights to optimize the metrics for specific policy objectives. Second, we present a novel method to update risk thresholds over time and show that this adaptive approach outperforms static metrics. With our approach, we demonstrate that metrics using only new hospital admissions perform as well in prediction as metrics that also include cases and inpatient bed usage.

## Materials and Methods

The CDC used indicators available nationwide (cases, hospitalizations, and occupancy of staffed inpatient beds) to develop Community Levels (2). In this research, we used the same indicators to define alternative state and county metrics, then compared these metrics based on ability to predict future health outcomes.

### Outcomes

The primary evaluation criterion was predictive power for high mortality. We defined “high mortality” as >1 death per 100,000 per week and “very high mortality” as >2 deaths per 100,000 per week. The lower threshold was defined in reference to peak mortality of other respiratory viruses (influenza and respiratory syncytial virus) during a severe season (7, 14). Let *T*_*M*_ *∈*1, 2 denote these mortality thresholds. The true outcome was a binary variable equal to 1 if mortality three weeks from the current week (i.e., at time *w* + 3) in location *i* exceeded the threshold; formally, *Y*_*i,w*+3_ = I(mortality at *w* + 3 *> T*_*M*_) *∈* 0, 1. In secondary analyses, we evaluated predictive power for ICU admissions, for which we defined “high” as >2 prevalent ICU admissions per 100,000 population per week.

We used a 3-week prediction window because previous CDC analyses indicated that this maximized the correlation between indicators and outcomes (2). This also reflects the necessary lead-time for interventions to have an impact on severe outcomes; a metric that predicts severe mortality tomorrow will come too late for effective action. We used binary outcomes to mirror CDC risk categories and to reflect the common practice of adopting pandemic response interventions in response to threshold crossing.

### Indicators

Indicators are the observed quantities that enter our prediction models. We used the same three indicators as the CDC’s Community Levels: new COVID-19 cases per 100,000 (weekly total), new COVID-19 hospital admissions per 100,000 (weekly total), and the occupancy of staffed inpatient hospital beds by COVID-19 patients (7-day average). Let *X*_*C,i,w*_, *X*_*H,i,w*_, and *X*_*O,i,w*_ denote the levels of these three indicators respectively, in location *i* during week *w*. In our risk prediction models, we used these indicators in 5 combinations: 1) new cases only (C), 2) new hospital admissions only (H), cases and hospital admissions (CH), 4) hospital admissions and bed occupancy (HO) and 5) all three indicators (CHO).

### Data

We obtained data on indicators and outcomes at both state and county levels and conducted separate analyses for each geographic level. For cases and deaths, we used aggregated counts compiled by state and local health agencies (15). For new COVID-19 admissions and bed occupancy, we used data reported to the U.S. Department of Health and Human Services Unified Hospital Data Surveillance System (16, 17). Consistent with CDC Community Level calculations, we calculated county-level hospitalizations at the Health Service Area-level to account for care-seeking across counties and computed measures at the midpoint of each week (2). (HSAs were defined by the National Center for Health Statistics to be one or more contiguous counties with self-contained hospital care (18).) In sensitivity analyses, we also present analyses with all inputs and outcomes calculated at the HSA-level.

### Metrics

Metrics take indicators as inputs and produce a binary high risk classification for a geographic area as output. Our metrics used data available at week *w* to predict mortality above the pre-specified threshold for mortality, *T*_*M*_ three weeks in the future and then classify a locality as high risk, Ŷ _*w*+3_ = 1, or not high-risk Ŷ _*w*+3_ = 0. (For readability, we omit location subscripts *i* when referring to a single observation in this section.)

### Objective

We used weighted classification accuracy to compare metrics on their ability to predict future high mortality, where the weights reflect preferences for avoiding different types of errors.

We assumed a simple underlying decision-analytic framework: a decision maker receives a prediction of mortality three weeks hence, *Ŷ*_*w*+3_, and takes action in response to that prediction. If the metric predicts high mortality (*Ŷ*_*w*+3_ = 1), she will take one action; if the model does not predict high mortality (*Ŷ*_*w*+3_ = 0), she will take a different action. Each action has benefits and costs that depend on the true outcome. For example, a true negative conserves public health resources, while a false negative may have costs such as failing to prevent a hospital from becoming overburdened. By contrast, a false positive may have costs such as wasted resources and harming public trust due to unnecessary policy actions.

We consider costs in terms of disease burden and public health resources. We anchor costs at 0 in the scenario in which the model correctly predicts low mortality (Ŷ_*w*+3_ = *Y*_*w*+3_ = 0). If the model incorrectly predicts high mortality (Ŷ_*w*+3_ = 1, *Y*_*w*+3_ = 0), we denote costs *R*_0_, of public health resources spent and social costs. By contrast, if a model incorrectly predicts low mortality (Ŷ_*w*+3_ = 0, *Y*_*w*+3_ = 1), policymakers incur a cost of *D*, of disease. Last, if a model correctly predicts high mortality (Ŷ_*w*+3_ = *Y*_*w*+3_ = 1), we assume policymakers implement an intervention that reduces disease by a factor of *α*, but pay resource costs, for a total cost of (1*− α*)*D* + *R*_1_.

The total cost associated with a particular metric (omitting subscripts for parsimony) is:

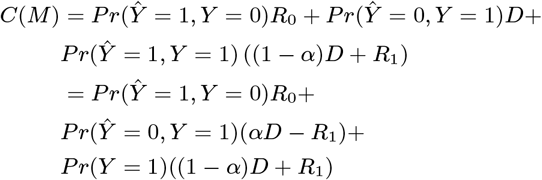

Because the last term is constant across all metrics (which cannot affect prevalence of high risk states), this cost is proportional to the weighted misclassification rate:

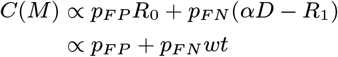

We can therefore rank metrics based only on performance (i.e., their probabilities of making each error type) and the decision maker’s relative preference for false positives compared to false negatives (*wt*). As the above expression indicates, we can conceptualize weight *wt* as the ratio of the net benefit from taking action on a true positive (*αD − R*_1_) to costs incurred by unnecessary action in the case of a false positive (*R*_0_).

In our primary analyses, we considered three values of this weight: “neutral” weighted false negatives and false positives equally (*wt* = 1, equivalent to unweighted accuracy), “don’t cry wolf” down-weighted false negatives as half the cost of false positives (*wt* = 0.5), and “better safe than sorry” downweighted false positives as half the cost of false negatives (*wt* = 2).

We estimated the weighted accuracy rate for each metric as 1 minus the weighted misclassification rate:

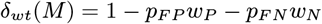

While any *w*_*N*_ and *w*_*P*_ such that 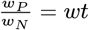 would produce the same ranking of metrics, the absolute value of *δ*_*wt*_ depends on these *w*_*N*_ and *w*_*P*_, which compare the cost of errors to the benefits of a correct classification. We set *w*_*N*_ and *w*_*P*_ such that both error weights are shifted equally in magnitude to achieve the desired ratio, with an increase in one and corresponding decrease in the other. That is, we set *w*_*N*_ and *w*_*P*_ using the value *a* such that *w*_*N*_ */w*_*P*_ = (1*− a*)*/*(1+*a*) = *wt*. With neutral weighting, *w*_*N*_ = *w*_*P*_ = 1.

We used weighted accuracy as our primary measure of performance, with higher weighted accuracy indicating better performance. We further weighted *δ*_*wt*_ by population to reflect the total proportion of individuals living in a location with an accurate classification (SI Text A).

### Static metrics

We considered two types of metrics, static and adaptive. Static metrics used the same procedure in each period to classify a locality as high risk. They differed in their input indicators (the sets C, H, CH, HO, and CHO described above) and the corresponding thresholds used to classify a locality as high risk. We varied the threshold on cases from 0 to 300 per 100,000 (in increments of 50), on new hospitalizations from 0 to 25 per 100,000 (in increments of 5), and on occupancy from 0 to 20% (in increments of 5). In what follows, let *T*_*C*_ *∈* [0, 300], *T*_*H*_ *∈* [0, 25], and *T*_*O*_ *∈* [0, 20] denote the thresholds for cases, hospitalizations, and occupancy, respectively. We designated the area as high risk if all the indicators in a given indicator set are above their specified thresholds.

We also replicated the CDC’s Community Levels, designating an area as high-risk if

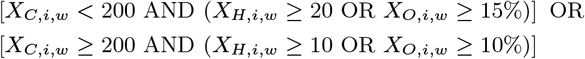

Last, we considered a metric (Z) that designates an area as “high risk” if the outcome is currently above the threshold of interest, i.e. *Ŷ*_*i,w*+3_ = I (*Y*_*i,w*_ = 1).

### Adaptive metrics

Adaptive metrics changed thresholds over time based on their ability to predict mortality during the recent past (Figure 1). At time *w*, we used as training data recent weeks of past indicator data with complete information on outcomes 3 weeks in the future. To these training data, we fit logistic regression models with outcomes on the left-hand side and indicators from three weeks previous on the right-hand side. For example, in the model corresponding to the CHO indicator set, we fit

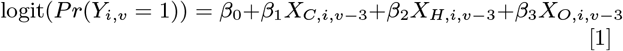

for *v* [*w* 3, *w*]. From this model, we obtained 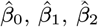, and 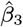, which we then used to produce fitted probabilities for each locality’s mortality three weeks ahead using:

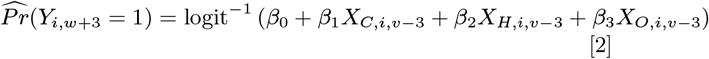

**Fig. 1.**
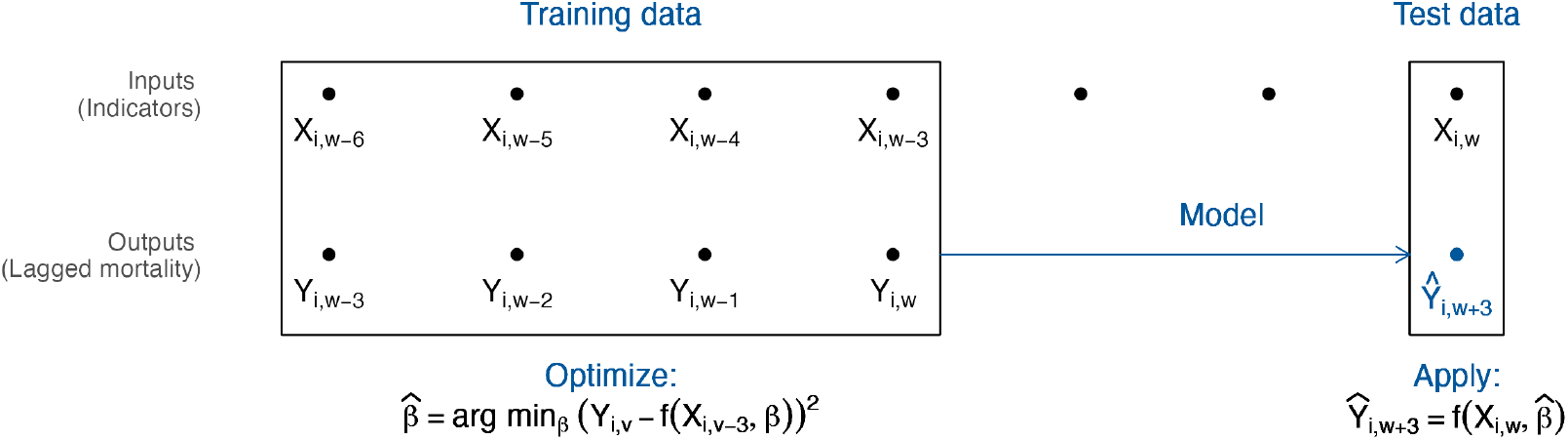
Adaptive metrics. We used input data from time *w* to predict mortality at time *w* + 3. The diagram shows the model-fitting process using 4 weeks of training data. We trained a model using the 4 most recent weeks with complete outcome data, including inputs from *w −* 6 to *w −* 3 and outputs from *w −* 3 to *w*. We then used this model, with input data from *w*, to estimate the probability of “high” or “very high” future mortality at *w* + 3 and designated a binary prediction based on whether this probability exceeded the user’s cutoff. (When a single indicator is used as the only input, this process is equivalent to identifying the optimal threshold for the indicator over the training period, accounting for user preferences.)

Logistic regression smoothed over noise in the small training data and reduced the dimension of multiple indicators by converting to a probability scale.

With predictions on a probability scale, we specified a probability cutoff above which we classified a location as high risk. We selected this cutoff based on the relative weighting of different error types (*wt*). We classified a locality as high risk whenever the probability was above 1*/*(1 + *wt*) (see SI Text B for optimal cutoff derivation). For our three weights (neutral, don’t cry wolf, and better safe than sorry), the cutoff values were 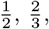, and 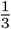, respectively. With a single predictor, this process would be equivalent to identifying the optimal threshold for the indicator over the training period, accounting for user preferences.

We specified analogous models based on CHOZ and HZ indicator sets to assess sensitivity to different functional forms. We also included a simplified version that was updated less frequently, only re-fitting to the training data each quarter, rather than each week. We varied the number of training weeks from 3 to 12 (i.e., fitting Eq. 1 to training data sets as large as *v ∈* [*w −* 11, *w*]).

### Head-to-head comparison

We compared the performance of the metrics during a training period. To define the training period, we began with the period the CDC used to fit Community Levels (March 1, 2021 through January 24, 2022). We further allowed the month of March for model fitting and including 3 weeks of future mortality data. Thus, our training period covered April 1, 2021 through December 31, 2021, that is, 2021 Q3 and Q4, with outcomes extending through January 21, 2022.

We compared performance across metrics separately for each outcome (*>* 1 or *>* 2 deaths/100k/week and >2 ICU admissions/100k/week), preference weight (*wt* = 0.5, 1, or 2), and geographic area (state or county). Within each combination of these, we chose the best-performing static metric during the training period from among the 7, 6, 42, 24, or 168 possibilities within the C, H, CH, HO, and CHO indicator sets and for adaptive metrics, we selected the best performing number of training weeks. The CDC Community Levels and current outcome (Z) metrics were fixed, so there was no selection within this metric type.

### Performance evaluation

We present weighted accuracy of each selected metric in the training quarters (during which the best performer of each type was selected) and a test period of January 1, 2022 through September 30, 2022 (i.e., 2022 Q1-Q3). As a sensitivity analysis, we used December 15, 2021 through February 15, 2022 as a training period, to include only omicron-specific training data, and data from February 16 through September 20, 2022 as test data.

In addition to presenting overall weighted accuracy, we summarize variation in performance across quarters with maximum quarterly regret, the difference between a metric’s predictive accuracy and the best performing metric (19). We calculate regret for each selected metric in each quarter and take the maximum across quarters:

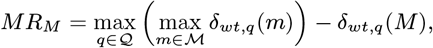

where *M* is a metric of interest, Q is a set of quarters, M is a set of metrics, and *δ*_*wt,q*_ is weighted accuracy during quarter *q*.

Last, to decompose variation between metrics into differences in predictive power and differences in error preferences, we computed sensitivity (*Pr*(*Ŷ*_*i,w*+3_= 1|*Y*_*i,w*+3_ = 1)) and specificity (*Pr*(*Ŷ*_*i,w*+3_ = 0|*Y*_*i,w*+3_ = 0)) across different *wt* values for adaptive metrics and compared these to sensitivity and specificity for static metrics.

### Simulations

To generalize our approach beyond the specific pandemic periods considered, we developed simple simulations, varying the change in relationship between indicators and outcomes over time and indicator distribution/prevalence of “high” outcomes (SI Text C). We then estimated predictive accuracy across different scenarios.

## Results

Indicator levels and lagged mortality varied substantially over the course of the study period (Figure 2), which included two major waves of high mortality (delta and omicron BA.1) and a smaller wave in summer 2022 (omicron BA.5) (See Figures S2-S3 for detailed dynamics of indicators by outcome over the study period.) The percentage of population-weighted state-weeks with high lagged mortality ranged from 94% during Q4 2021 to a low of 17% during Q2 2021. For very high mortality, this ranged from 61% (Q1 2022) to 3% (Q2 2022). We observed similar variation in counties, with less extreme swings (e.g, from 25% to 75% for high mortality). The relationship between indicators and outcomes shifted substantially over the period studied. In particular, in the third quarter of 2022, cases, hospitalizations, and bed occupancy all increased, but mortality remained lower than in previous waves (Figure 2).

**Fig. 2.**
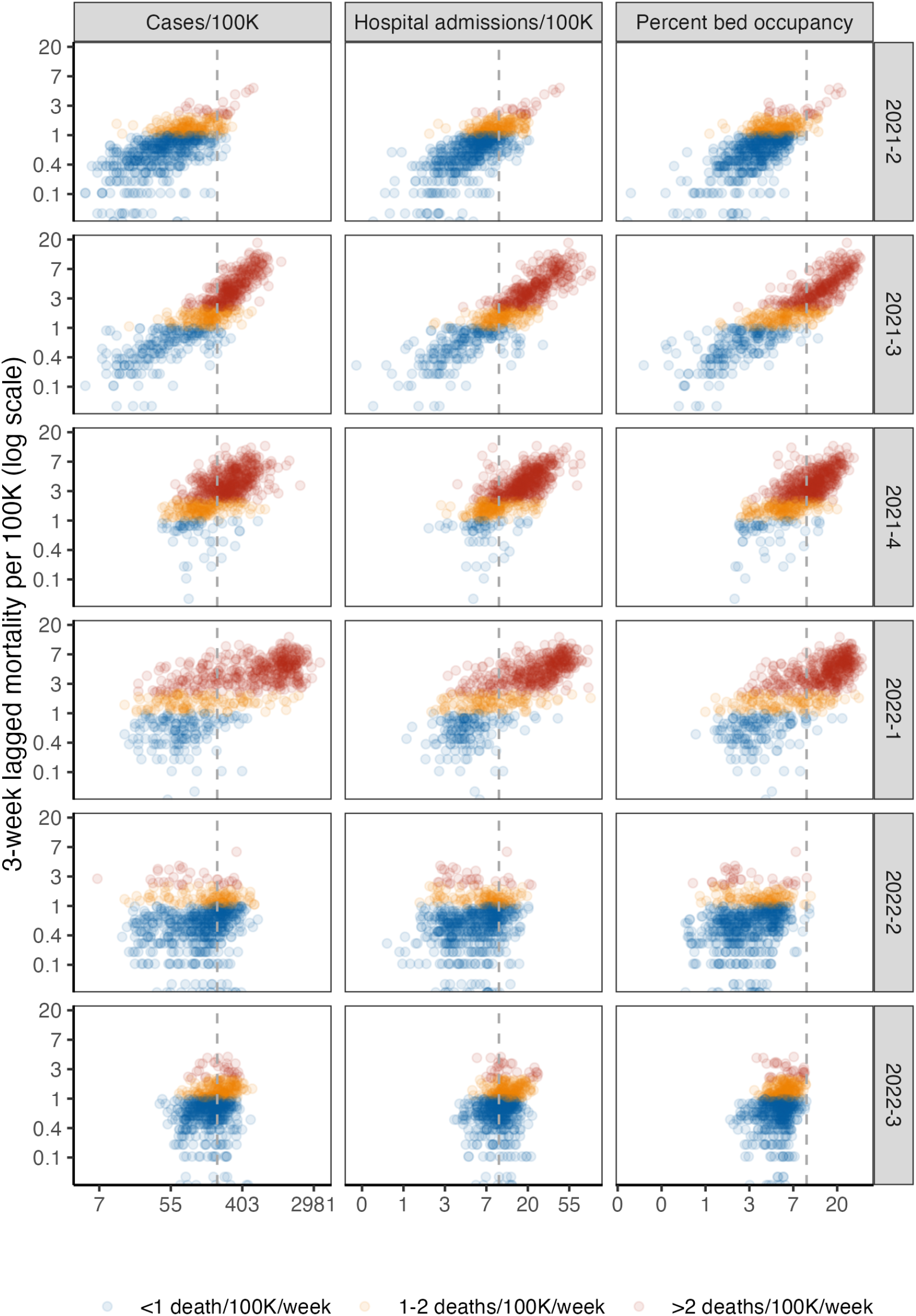
State-level lagged mortality vs. indicator levels by quarter. Columns indicate different indicators (weekly cases per 100,000 population, new hospital admissions per 100,000, and percentage of inpatient beds occupied by COVID-19 patients), and rows indicate quarters. The x-axis displays indicator values on a log scale and y-axis displays 3-week ahead mortality per 100,000 population on a log scale. Each point on the scatterplot is a state-week. Colors show mortality outcome level. The vertical gray dotted lines indicate thresholds from CDC Community Levels for each indicator (*≥* 200 cases/100K/week and *≥* 10 new admissions/100K/week or *≥* 10% COVID-19 bed occupancy.) See Figure S1 for a county-level plot.

### Static metrics

In Figure 3, we present the performance of the best-performing static metrics from different indicator sets (C, H, CH, HO, and CHO) during the training and test periods. During the training period, there were only minor differences in training accuracy between metrics that used different indicator sets (e.g., 83-87% in predicting high mortality for states with neutral weighting, 73%-75% for counties). However, for nearly all static metrics and outcomes, test accuracy was lower and more variable than training accuracy (e.g., 45-68% and 54-72% for high mortality in states and counties respectively).

**Fig. 3.**
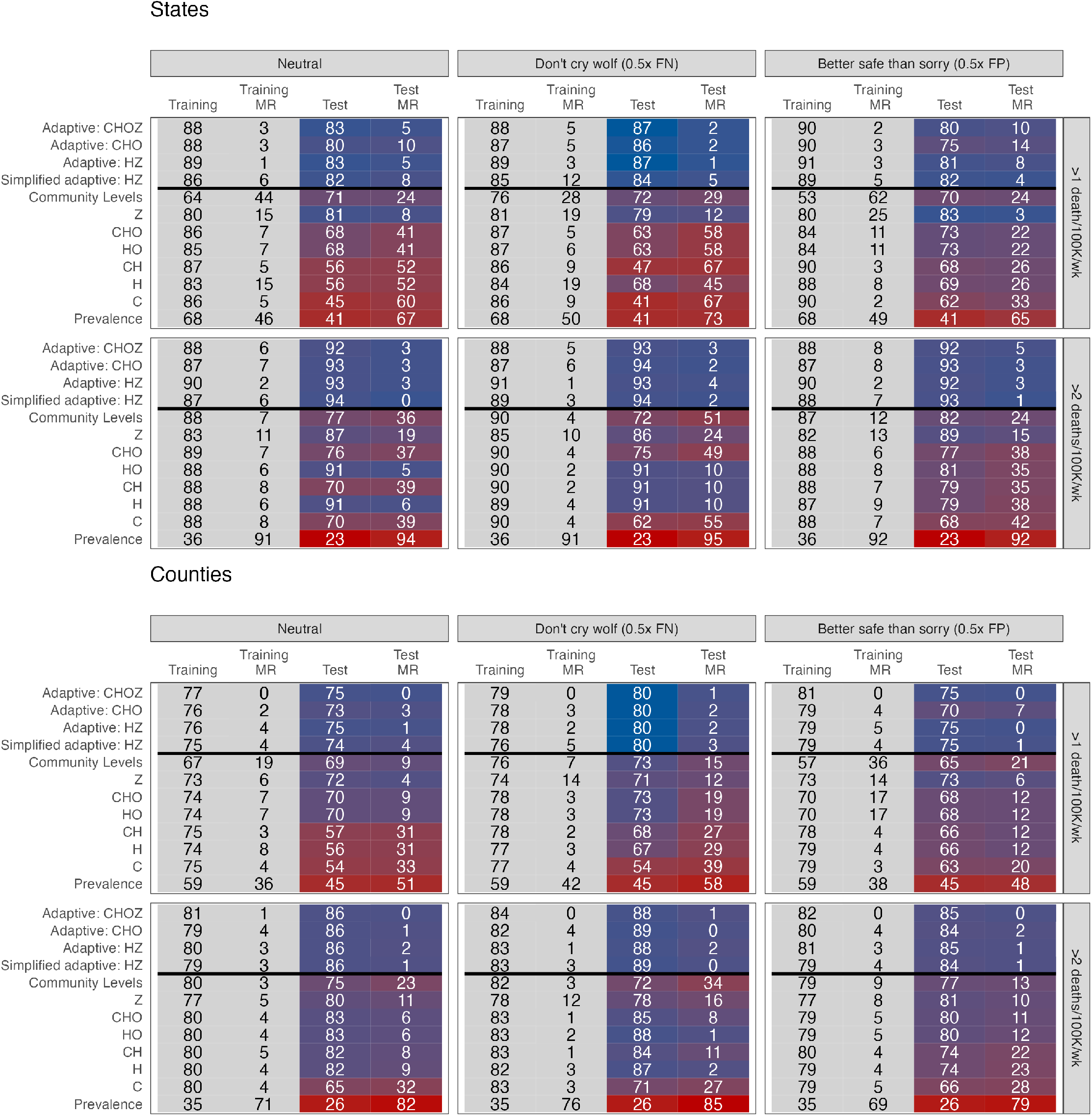
Head-to-head comparison results. The top plots display results from state-level analyses and the bottom plots display results from county-level analyses, both weighted for population. Metrics are displayed on the left, with training data from Q2-Q4 2021 and test data from Q1-Q3 2022. Cells report weighted accuracy and maximum regret (MR) over training and test periods. Rows vary outcomes, and columns vary preferences for false positive versus false negatives, with “neutral” corresponding to unweighted accuracy. Prevalence indicates the proportion of high location-weeks in a given time period. A version including HSA-level analyses can be found in Figure S4. Weighted accuracy by quarter, including for intensive care usage, is presented in Figures S5-S7.

Some of this variation is due to the shifting relationship between indicators and lagged outcomes over time. We illustrate this in Figure 4, where gray lines show the performance of metrics based on different hospitalization cutoffs. No single cutoff dominated during the full study period. For example, the cutoff of 5 per 100,000 performed best for high mortality during the first 3 quarters of the study period, with accuracy above 90% in states and 74% in counties, but was the worst performing in Q2-Q3 2022, with less than 50% accuracy. The accuracy of the single best-performing metric also varied across quarters (e.g., from 68-81% for high mortality and 79-91% for very high mortality in counties).

**Fig. 4.**
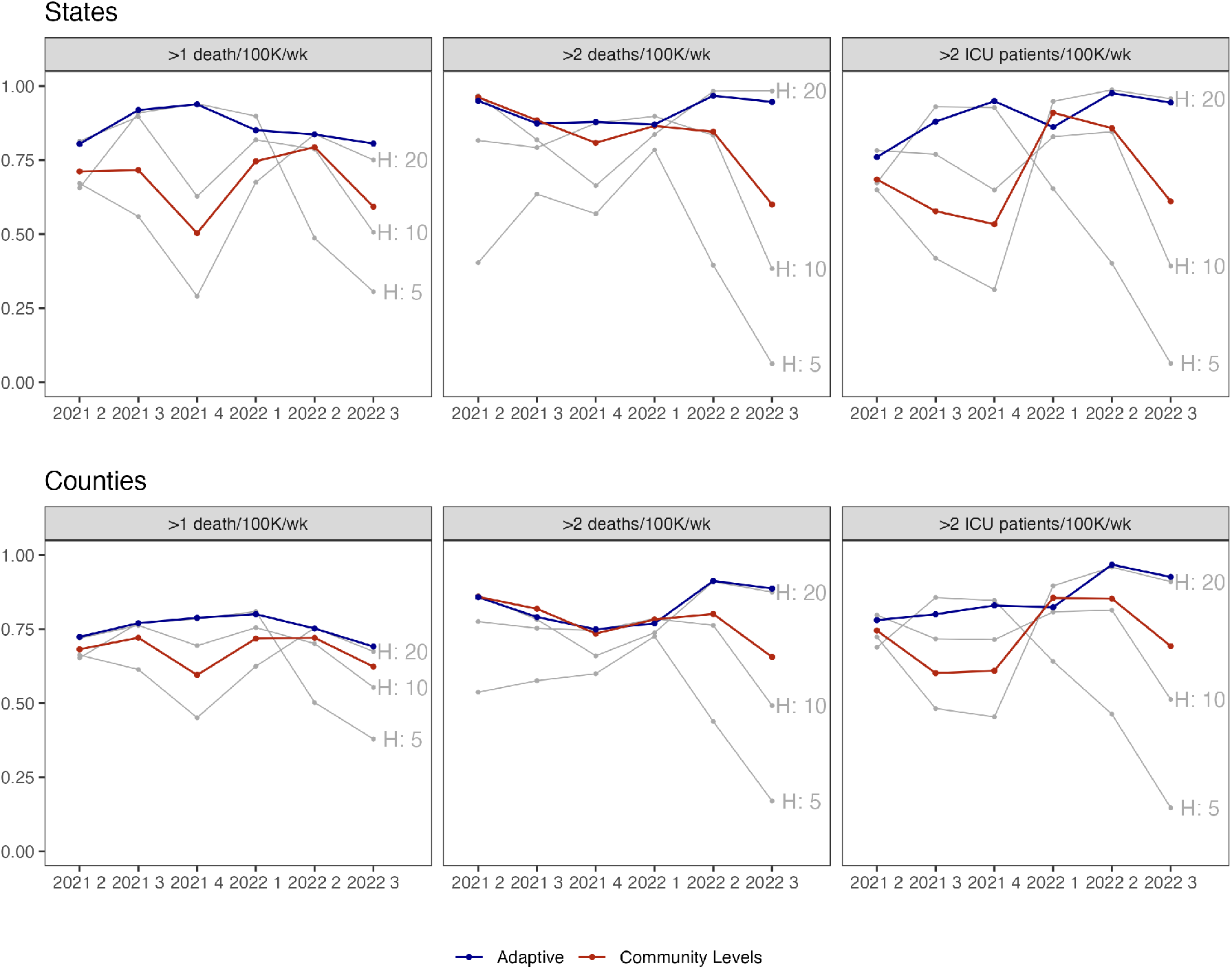
Weighted accuracy by metric. The top plot displays states, and the bottom plot displays counties. Columns indicate different outcomes. The x-axis indicates quarter, and the y-axis predictive accuracy (neutral weighting). Grey lines depict metrics based on new hospital admissions exceeding the row threshold. The red line indicates CDC Community Level and the blue line an adaptive metric (HZ). A version with HSA-level results can be found in Figure S8.

Other static metrics similarly reflected the evolving relationship between indicators and mortality. While prediction based on current outcome (Z) was the second-worst performing static indicator during the training period (after Community Levels), it performed best during the test period, when waves of infection were less extreme and variable. CDC Community Levels performed relatively worse compared to other static metrics at predicting high mortality during the training period, but similar or better during the test period; the converse was true for predicting very high mortality (Figure 3). Overall, metrics that used hospitalizations and bed occupancy performed most consistently across training and test periods, but we would have been unable to discern this with only training data. Across static metrics, training accuracy was an unreliable signal of test accuracy.

### Adaptive metrics

Adaptive metrics consistently outperformed static metrics for both outcomes in training and test periods (Figure 3). For example, when predicting high mortality in states with neutral weighting, adaptive metrics had overall accuracy of 86-89% in the training period and 80-83% in the test period; for very high mortality, this was 87-90% and 92-94% respectively. While all adaptive functional forms performed well, metrics corresponding to CHOZ and HZ slightly outperformed CHO and the simplified version with less frequent updating. Importantly, while adaptive metrics performed similarly to static metrics during some quarters, they rarely underperformed by a substantial margin and often achieved substantial gains (Figure 4). This was reflected in regret, which was minimized by CHOZ and HZ adaptive metrics for both outcomes. CHOZ and HZ adaptive metrics also weakly dominated static indicator-based metrics and Community Levels in the sense that they could achieve at least equal (and often higher) sensitivity and specificity for at least one value of *wt* (Figure S9).

### Alternative preferences, secondary outcomes, and sensitivity analyses

Adaptive metrics similarly outperformed static metrics for across preference weights (Figure 3) and for a secondary outcome of ICU bed usage over 2 per 100,000 (Figure 4). The gain in weighted accuracy for adaptive metrics was higher when estimated at the HSA level rather than at the county level (about 2 percentage points for both mortality outcomes with neutral weighting). Running the training period from December 15 to February 15 to capture the omicron variant did not substantially alter the relative benefit of adaptive metrics, with a 14 percentage point increase in weighted accuracy in states for high mortality compared to Community Levels with a neutral weighting (compared to 11% in the base case) and 7% in counties (compared to 6%).

### Simulations

In simulations, adaptive methods outperformed static methods when the relationship between indicators and outcomes was changing over time, regardless of whether outcome prevalence was constant or wave-driven. There was no gain when the relationship between indicators and outcomes was static; adaptive metrics performed worse than static metrics when indicator prevalence was highly variable, and there could be insufficient training data near the threshold to estimate the optimal cutoff.

## Discussion

We proposed an adaptive approach to estimating local risk which continually updates metrics to ensure they predict outcomes of policy interest. We showed that this would have outperformed static approaches, including CDC Community Levels over the past year. Our metrics have a unique advantage in a rapidly evolving pandemic context. They quickly pick up new information as the relationship between indicators and lagged mortality shifts, allowing us refine the threshold for “high risk” and improve discrimination.

Previous papers have proposed adaptive policies for COVID-19 management, in which policymakers shift responses depending on observed indicators like cases and deaths (20–22). We extend this work by allowing the trigger thresholds for indicators to also vary over time. Such an approach could be particularly advantageous for maintaining public trust when the relationship between indicators and outcomes is not yet well-understood or is changing over time (23).

Our approach draws on ideas that have been applied in the online calibration literature and in forecasting, but have not yet been widely applied for population risk metrics (6, 24– 26). In contrast to some other applications, we particularly emphasize parsimony for policy metrics, demonstrating that policymakers can obtain equal predictive performance with fewer input indicators, potentially reducing the burden of data collection on state and local public health departments. Similar to other authors, we find hospitalizations to be a particularly powerful predictor of future mortality (6). We further emphasize that it is valuable to collect real-time data on outcomes of policy interest, like mortality. (In the case of COVID-19, while state mortality is still collected and reported weekly, many counties have reduced reporting frequency (15).)

Our method can also reflect a policymaker’s preferences for the trade-off between avoiding false negative and false positives, filling a previously-identified gap between models and decision theory (27). In practice, different indicators could be used to guide different policies. For the most burdensome policies (e.g., business closures), policymakers might prefer a low risk of false negatives, while less burdensome policies (e.g., distribution of rapid tests) might have a higher tolerance for false positives.

There are several limitations to this study. First, we model only outcomes related to severe disease and death from COVID19, as national policymakers have designated these priority outcomes. Nevertheless, metrics to track illness are also important for understanding the full burden of COVID-19, which can also include disruptions from illness as well as Long COVID, as is work to predict surges with longer lead time (26, 28). In addition, no adaptive framework can automatically incorporate all possible variation. Manual tuning may be needed, for example, if the frequency of reporting of hospitalization changes over time. Furthermore, in high-risk situations, such as an unusually lethal new variant identified in one country, it may be preferable to implement preventative measures even prior to observing a changing relationship between indicators and severe outcomes. More broadly, metrics could be refined to upweight performance during critical periods such as the start of a surge or consider dynamic decision-making. Finally, future work could also expand these results to other contexts, such as prediction of combined respiratory disease outcomes (including influenza and RSV) and consider other models for risk prediction. Overall, adaptive metrics may be a powerful tool for designing trustworthy, transparent metrics to guide infectious disease policy.

## Data Availability

Data and code are publicly available on GitHub.

https://github.com/abilinski/AdaptiveRiskMetrics

## Data Archival

Data and code will be made publicly available on GitHub.

## ACKNOWLEDGMENTS

The authors were supported by the Centers for Disease Control and Prevention though the Council of State and Territorial Epidemiologists (NU38OT000297-02; AB, JAS) and the National Institute on Drug Abuse (3R37DA01561217S1; JAS). Funders had no role in the design and conduct of the study; collection, management, analysis, and interpretation of the data; preparation, review, or approval of the manuscript; and decision to submit the manuscript for publication.

## Notes

### Competing Interest Statement

The authors have declared no competing interest.

