## Supplement for "Adaptive metrics for an evolving pandemic A dynamic approach to area-level COVID-19 risk designations"

**This PDF file includes:**

Supporting text
Figs. S1 to S12

### Supporting Information Text

**A. Population-Weighted Performance.** Let $S_i$ be the population of location $i$. The weight for location $i$ is $s_i = \frac{S_i}{\sum_{i=1}^{N} S_i}$. To obtain population-weighted estimates of our accuracy measure, we estimate:

$$\delta_{wt} = 1 - \sum_i s_i \left( p_{FP} w_P + p_{FN} w_N \right)$$

**B. Optimal Threshold Selection.** We first specify a weight $wt$ such that we consider false positives to be a factor of $wt$ as costly as false negatives. For example, if $wt = 3$, we consider 3 false positives as costly as 1 false negative; 2 false positives would be less costly than 1 false negative. Denote the probability that an outcome of interest occurs given observed indicators for an observation, $q_{i,w+3} = Pr(Y_{i,w+3} = 1|X_i)$. We want to predict that the outcome will occur if, in expectation, this will decrease net costs. If an observation with probability $q_{i,w+3}$ is classified with a prediction of 0, this has probability $q_{i,w+3}$ of being a false negative. If it is classified with a prediction of 1, there is a probability $1 - q_{i,w+3}$ of a false positive. We therefore should classify with a prediction of 1 if:

$$\begin{aligned} \text{Expected cost of FP} &\leq \text{Expected cost of FN} \\ (1 - q_{i,t+3}) &\leq q_{i,t+3} wt \\ q_{i,t+3} &\geq \frac{1}{1 + wt} \end{aligned}$$

**C. Simulations.** To conduct simulations, we generate data that assumes a logistic relationship between the probability of a high outcome ($Pr(Y_{i,w+3})$) and a synthetic hospitalization indicator ($X_{H,i,w}$):

$$logit\left(Pr(Y_{i,w+3} = 1|\, X_{H,i,w}\right) = \beta_0 + \beta_1 X_{H,i,w} + \epsilon_{iw}, \quad [1]$$

where $\epsilon_{iw}$ are $i.i.d$ draws from a logistic distribution with mean 0 and scale parameter $\sigma$. We then draw $Y_{i,w+3}$ from a binomial distribution with the corresponding probability.

We vary simulations across 3 main dimensions:

1. **Indicator prevalence:** We first use empirical hospitalization data for simulations, drawing synthetic outcomes according to 1. To build intuition, we then use two stylized scenarios, one in which prevalence is constant over quarters and one in which waves are even more pronounced.
   (a) Empirical: We use true state-level hospitalization data from Q3 2021 through Q4 2022.
   (b) Constant: We draw $X_{H,i,w}$ from a $Unif(2, 20)$ distribution for state-times from Q3 2021 through Q4 2022.
   (c) Sharp waves: We alternate each quarter between drawing hospitalizations from a $N(5, 1)$ distribution and a $N(15, 1)$ distribution for each state-time from Q3 2021 through Q4 2022.
2. **Relationship between inputs and outputs:** The optimal cutoff for a metric with neutral weighting is $-\beta_0/\beta_1$. We vary this as displayed in Figure S11:
   (a) Constant: 10 hospitalizations per 100,000 population
   (b) Linear increase: linearly increasing from 5 to 15 hospitalizations per 100,000 over the study period
   (c) Logistic increase: increasing from 5 to 15 hospitalizations per 100,000 over the study period per a logistic model with a sharp increase at week 25
   (d) Non-monotonic: optimal cutoff increases and then decreases

For illustration, we set $\beta_1 = 3, \sigma = 1$ and use the first quarter (synthetic Q3 2021) as training data. For each scenario, we simulate 50 draws. As in the main text, we select the best-performing static metric during training data and compare performance in terms of predictive accuracy to adaptive metrics, averaging over draws. Results are displayed in Figure S12.

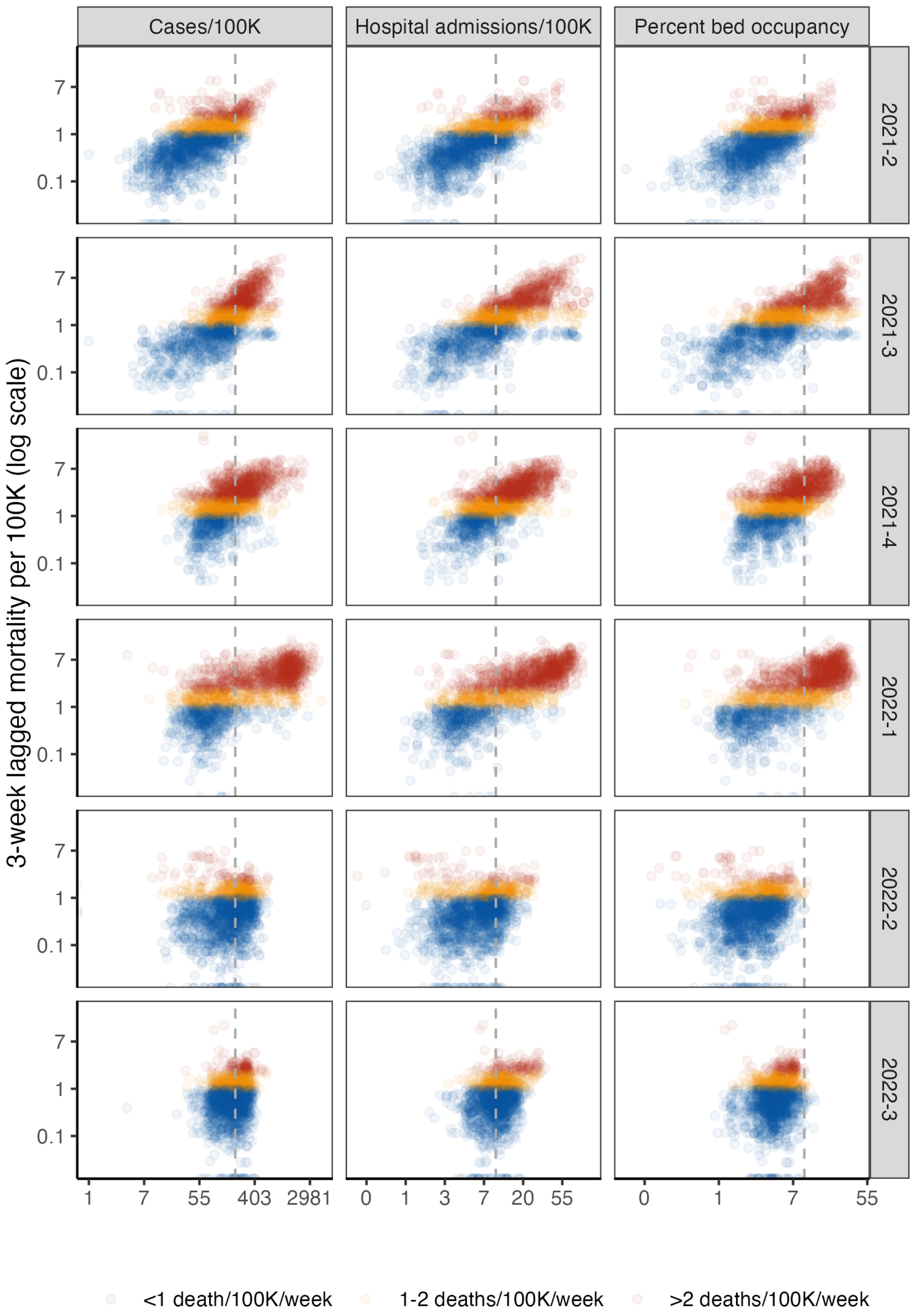


**Fig. S1.** County-level lagged mortality vs. indicator levels by quarter. Columns indicate different indicators (weekly cases per 100,000 population, new hospital admissions per 100,000, and percentage of inpatient beds occupied by COVID-19 patients), and rows indicate quarters. The x-axis displays indicator values on a log scale and y-axis displays 3-week ahead mortality per 100,000 population on a log scale. Each point on the scatterplot is a county-week. Colors show mortality outcome level. The vertical gray dotted lines indicate thresholds from CDC Community Levels for each indicator ($\geq 200$ cases/100K/week and $\geq 10$ new admissions/100K/week or $\geq 10\%$ COVID-19 bed occupancy.)

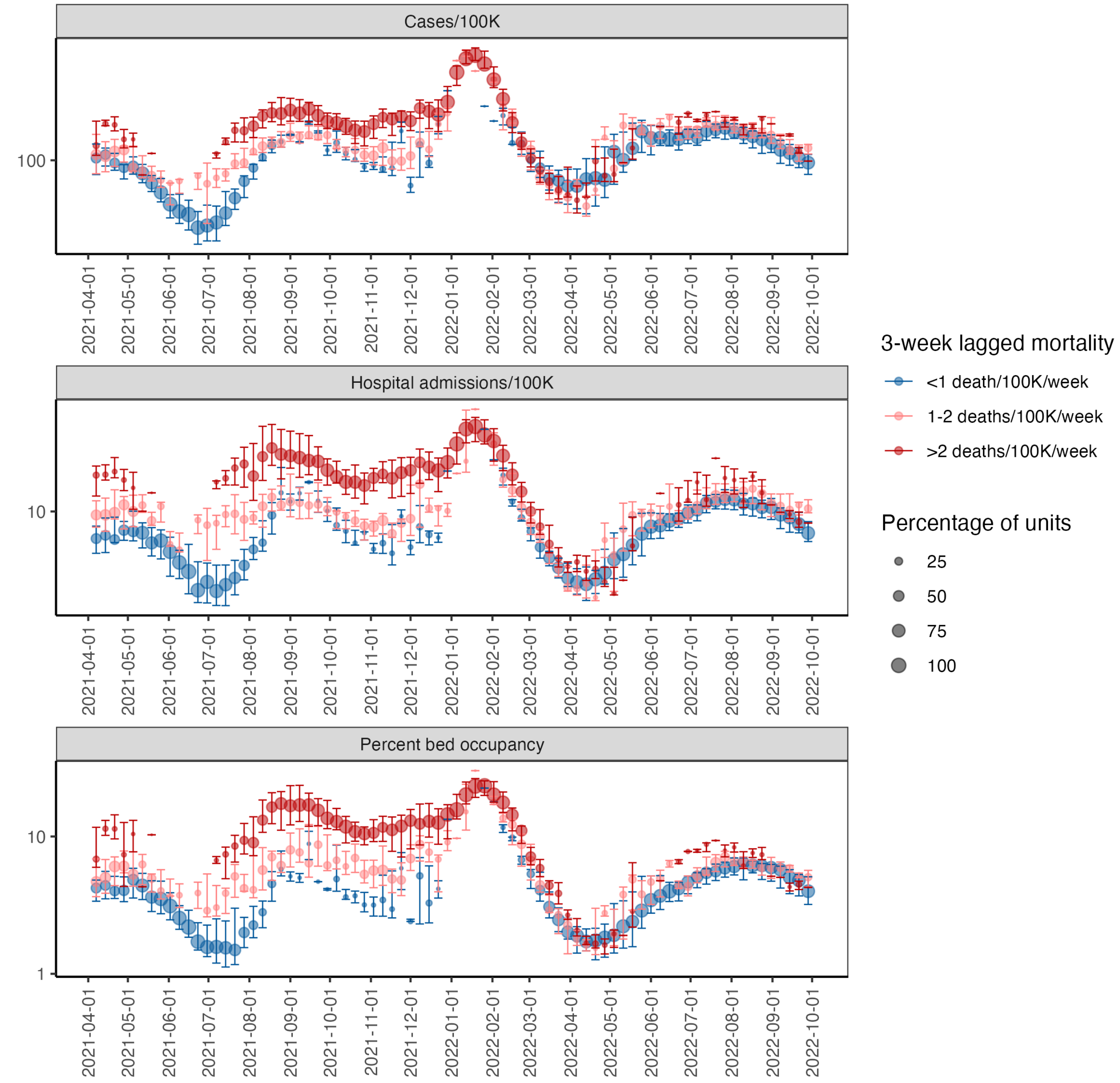


**Fig. S2.** Indicators by lagged mortality (state). Indicators vary across rows. The x-axis displays time and the y-axis displays the median (point) and interquartile range (bars) of each indicator by future mortality status.

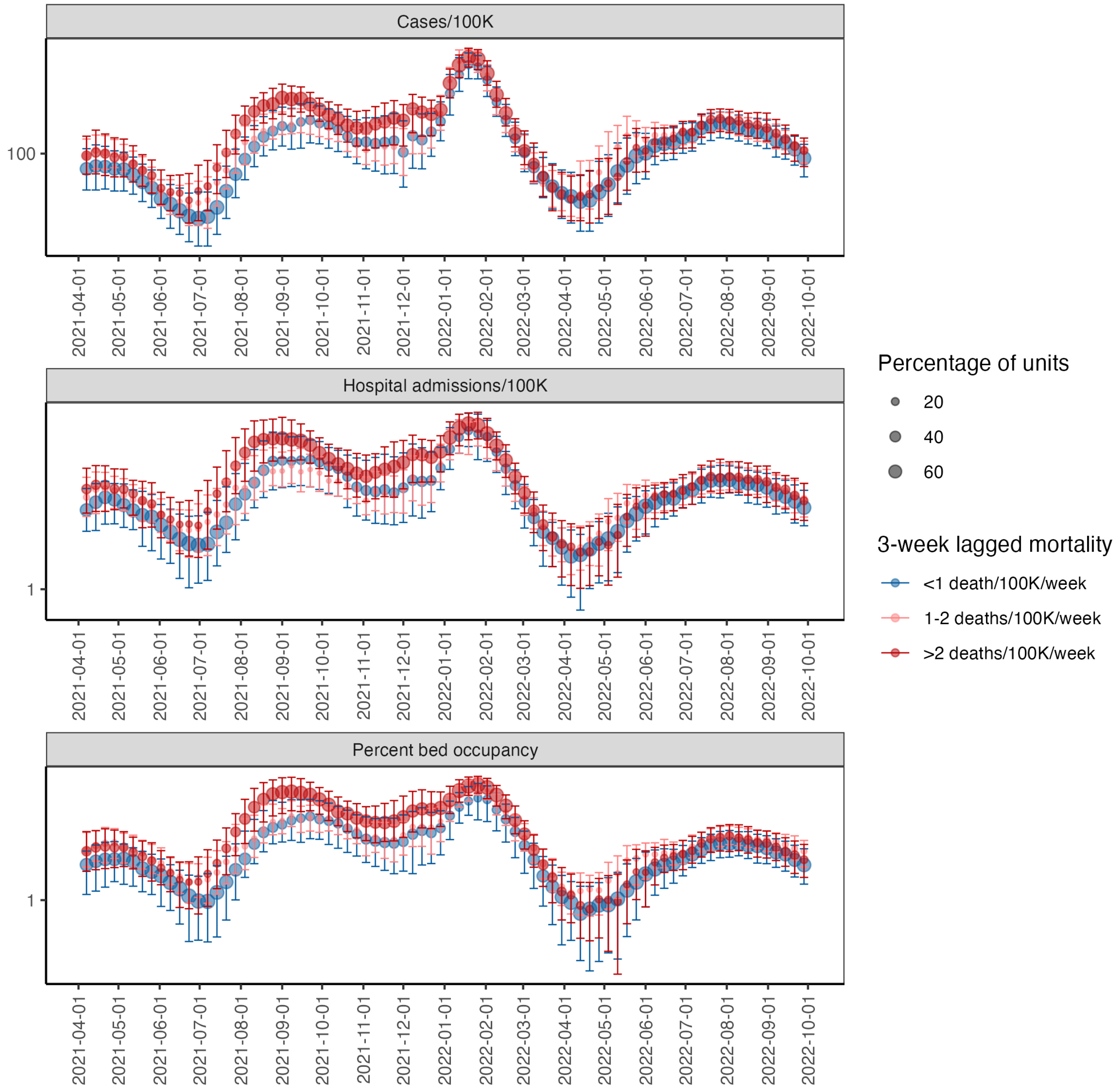


**Fig. S3.** Indicators by future mortality (county). Indicators vary across rows. The x-axis displays time and the y-axis displays the median (point) and interquartile range (bars) of each indicator by future mortality status.

### States

**>1 death/100K/wk**

| | Neutral: Training | Neutral: Training MR | Neutral: Test | Neutral: Test MR | Don't cry wolf (0.5x FN): Training | Don't cry wolf (0.5x FN): Training MR | Don't cry wolf (0.5x FN): Test | Don't cry wolf (0.5x FN): Test MR | Better safe than sorry (0.5x FP): Training | Better safe than sorry (0.5x FP): Training MR | Better safe than sorry (0.5x FP): Test | Better safe than sorry (0.5x FP): Test MR |
|---|---|---|---|---|---|---|---|---|---|---|---|---|
| Adaptive: CHOZ | 88 | 3 | 83 | 5 | 88 | 5 | 87 | 2 | 90 | 2 | 80 | 10 |
| Adaptive: CHO | 88 | 3 | 80 | 10 | 87 | 5 | 86 | 2 | 90 | 3 | 75 | 14 |
| Adaptive: HZ | 89 | 1 | 83 | 5 | 89 | 3 | 87 | 1 | 91 | 3 | 81 | 8 |
| Simplified adaptive: HZ | 86 | 6 | 82 | 8 | 85 | 12 | 84 | 5 | 89 | 5 | 82 | 4 |
| Community Levels | 64 | 44 | 71 | 24 | 76 | 28 | 72 | 29 | 53 | 62 | 70 | 24 |
| Z | 80 | 15 | 81 | 8 | 81 | 19 | 79 | 12 | 80 | 25 | 83 | 3 |
| CHO | 86 | 7 | 68 | 41 | 87 | 5 | 63 | 58 | 84 | 11 | 73 | 22 |
| HO | 85 | 7 | 68 | 41 | 87 | 6 | 63 | 58 | 84 | 11 | 73 | 22 |
| CH | 87 | 5 | 56 | 52 | 86 | 9 | 47 | 67 | 90 | 3 | 68 | 26 |
| H | 83 | 15 | 56 | 52 | 84 | 19 | 68 | 45 | 88 | 8 | 69 | 26 |
| C | 86 | 5 | 45 | 60 | 86 | 9 | 41 | 67 | 90 | 2 | 62 | 33 |
| Prevalence | 68 | 46 | 41 | 67 | 68 | 50 | 41 | 73 | 68 | 49 | 41 | 65 |

**>2 deaths/100K/wk**

| | Neutral: Training | Neutral: Training MR | Neutral: Test | Neutral: Test MR | Don't cry wolf (0.5x FN): Training | Don't cry wolf (0.5x FN): Training MR | Don't cry wolf (0.5x FN): Test | Don't cry wolf (0.5x FN): Test MR | Better safe than sorry (0.5x FP): Training | Better safe than sorry (0.5x FP): Training MR | Better safe than sorry (0.5x FP): Test | Better safe than sorry (0.5x FP): Test MR |
|---|---|---|---|---|---|---|---|---|---|---|---|---|
| Adaptive: CHOZ | 88 | 6 | 92 | 3 | 88 | 5 | 93 | 3 | 88 | 8 | 92 | 5 |
| Adaptive: CHO | 87 | 7 | 93 | 3 | 87 | 6 | 94 | 2 | 87 | 8 | 93 | 3 |
| Adaptive: HZ | 90 | 2 | 93 | 3 | 91 | 1 | 93 | 4 | 90 | 2 | 92 | 3 |
| Simplified adaptive: HZ | 87 | 6 | 94 | 0 | 89 | 3 | 94 | 2 | 88 | 7 | 93 | 1 |
| Community Levels | 88 | 7 | 77 | 36 | 90 | 4 | 72 | 51 | 87 | 12 | 82 | 24 |
| Z | 83 | 11 | 87 | 19 | 85 | 10 | 86 | 24 | 82 | 13 | 89 | 15 |
| CHO | 89 | 7 | 76 | 37 | 90 | 4 | 75 | 49 | 88 | 6 | 77 | 38 |
| HO | 88 | 6 | 91 | 5 | 90 | 2 | 91 | 10 | 88 | 8 | 81 | 35 |
| CH | 88 | 8 | 70 | 39 | 90 | 2 | 91 | 10 | 88 | 7 | 79 | 35 |
| H | 88 | 6 | 91 | 6 | 89 | 4 | 91 | 10 | 87 | 9 | 79 | 38 |
| C | 88 | 8 | 70 | 39 | 90 | 4 | 62 | 55 | 88 | 7 | 68 | 42 |
| Prevalence | 36 | 91 | 23 | 94 | 36 | 91 | 23 | 95 | 36 | 92 | 23 | 92 |

### HSAs

**>1 death/100K/wk**

| | Neutral: Training | Neutral: Training MR | Neutral: Test | Neutral: Test MR | Don't cry wolf (0.5x FN): Training | Don't cry wolf (0.5x FN): Training MR | Don't cry wolf (0.5x FN): Test | Don't cry wolf (0.5x FN): Test MR | Better safe than sorry (0.5x FP): Training | Better safe than sorry (0.5x FP): Training MR | Better safe than sorry (0.5x FP): Test | Better safe than sorry (0.5x FP): Test MR |
|---|---|---|---|---|---|---|---|---|---|---|---|---|
| Adaptive: CHOZ | 83 | 1 | 77 | 1 | 83 | 1 | 81 | 1 | 86 | 1 | 78 | 0 |
| Adaptive: CHO | 80 | 4 | 73 | 6 | 81 | 3 | 80 | 2 | 84 | 5 | 70 | 11 |
| Adaptive: HZ | 82 | 3 | 77 | 0 | 83 | 1 | 81 | 1 | 86 | 2 | 78 | 0 |
| Simplified adaptive: HZ | 81 | 3 | 76 | 1 | 80 | 7 | 80 | 3 | 85 | 3 | 78 | 1 |
| Community Levels | 64 | 31 | 68 | 13 | 75 | 14 | 73 | 13 | 52 | 49 | 63 | 27 |
| Z | 78 | 9 | 75 | 2 | 78 | 11 | 74 | 12 | 77 | 16 | 77 | 3 |
| CHO | 75 | 15 | 70 | 13 | 81 | 4 | 73 | 18 | 68 | 26 | 66 | 16 |
| HO | 75 | 15 | 70 | 13 | 81 | 4 | 73 | 18 | 68 | 26 | 66 | 16 |
| CH | 81 | 4 | 58 | 31 | 81 | 3 | 52 | 47 | 81 | 8 | 66 | 15 |
| H | 80 | 6 | 58 | 31 | 81 | 6 | 68 | 26 | 82 | 6 | 67 | 15 |
| C | 79 | 5 | 50 | 42 | 80 | 3 | 46 | 54 | 83 | 5 | 64 | 23 |
| Prevalence | 66 | 32 | 48 | 47 | 66 | 35 | 48 | 56 | 66 | 35 | 48 | 47 |

**>2 deaths/100K/wk**

| | Neutral: Training | Neutral: Training MR | Neutral: Test | Neutral: Test MR | Don't cry wolf (0.5x FN): Training | Don't cry wolf (0.5x FN): Training MR | Don't cry wolf (0.5x FN): Test | Don't cry wolf (0.5x FN): Test MR | Better safe than sorry (0.5x FP): Training | Better safe than sorry (0.5x FP): Training MR | Better safe than sorry (0.5x FP): Test | Better safe than sorry (0.5x FP): Test MR |
|---|---|---|---|---|---|---|---|---|---|---|---|---|
| Adaptive: CHOZ | 84 | 1 | 88 | 0 | 86 | 2 | 91 | 1 | 85 | 1 | 88 | 0 |
| Adaptive: CHO | 82 | 4 | 88 | 0 | 84 | 5 | 91 | 1 | 83 | 5 | 87 | 2 |
| Adaptive: HZ | 83 | 4 | 88 | 1 | 85 | 3 | 90 | 4 | 84 | 4 | 87 | 1 |
| Simplified adaptive: HZ | 82 | 5 | 87 | 2 | 83 | 5 | 91 | 1 | 82 | 4 | 86 | 2 |
| Community Levels | 83 | 5 | 76 | 24 | 85 | 2 | 73 | 37 | 81 | 13 | 78 | 14 |
| Z | 80 | 6 | 82 | 11 | 81 | 11 | 81 | 17 | 79 | 9 | 84 | 9 |
| CHO | 82 | 4 | 80 | 19 | 85 | 3 | 86 | 10 | 82 | 5 | 80 | 17 |
| HO | 82 | 7 | 85 | 5 | 85 | 2 | 86 | 10 | 82 | 7 | 81 | 12 |
| CH | 82 | 3 | 72 | 34 | 85 | 4 | 85 | 12 | 82 | 5 | 75 | 23 |
| H | 82 | 7 | 83 | 9 | 84 | 2 | 84 | 15 | 82 | 5 | 75 | 24 |
| C | 82 | 7 | 66 | 33 | 85 | 5 | 73 | 27 | 82 | 8 | 66 | 31 |
| Prevalence | 37 | 76 | 26 | 86 | 37 | 79 | 26 | 88 | 37 | 74 | 26 | 84 |

### Counties

**>1 death/100K/wk**

| | Neutral: Training | Neutral: Training MR | Neutral: Test | Neutral: Test MR | Don't cry wolf (0.5x FN): Training | Don't cry wolf (0.5x FN): Training MR | Don't cry wolf (0.5x FN): Test | Don't cry wolf (0.5x FN): Test MR | Better safe than sorry (0.5x FP): Training | Better safe than sorry (0.5x FP): Training MR | Better safe than sorry (0.5x FP): Test | Better safe than sorry (0.5x FP): Test MR |
|---|---|---|---|---|---|---|---|---|---|---|---|---|
| Adaptive: CHOZ | 77 | 0 | 75 | 0 | 79 | 0 | 80 | 1 | 81 | 0 | 75 | 0 |
| Adaptive: CHO | 76 | 2 | 73 | 3 | 78 | 3 | 80 | 2 | 79 | 4 | 70 | 7 |
| Adaptive: HZ | 76 | 4 | 75 | 1 | 78 | 2 | 80 | 2 | 79 | 5 | 75 | 0 |
| Simplified adaptive: HZ | 75 | 4 | 74 | 4 | 76 | 5 | 80 | 3 | 79 | 4 | 75 | 1 |
| Community Levels | 67 | 19 | 69 | 9 | 76 | 7 | 73 | 15 | 57 | 36 | 65 | 21 |
| Z | 73 | 6 | 72 | 4 | 74 | 14 | 71 | 12 | 73 | 14 | 73 | 6 |
| CHO | 74 | 7 | 70 | 9 | 78 | 3 | 73 | 19 | 70 | 17 | 68 | 12 |
| HO | 74 | 7 | 70 | 9 | 78 | 3 | 73 | 19 | 70 | 17 | 68 | 12 |
| CH | 75 | 3 | 57 | 31 | 78 | 2 | 68 | 27 | 78 | 4 | 66 | 12 |
| H | 74 | 8 | 56 | 31 | 77 | 3 | 67 | 29 | 79 | 4 | 66 | 12 |
| C | 75 | 4 | 54 | 33 | 77 | 4 | 54 | 39 | 79 | 3 | 63 | 20 |
| Prevalence | 59 | 36 | 45 | 51 | 59 | 42 | 45 | 58 | 59 | 38 | 45 | 48 |

**>2 deaths/100K/wk**

| | Neutral: Training | Neutral: Training MR | Neutral: Test | Neutral: Test MR | Don't cry wolf (0.5x FN): Training | Don't cry wolf (0.5x FN): Training MR | Don't cry wolf (0.5x FN): Test | Don't cry wolf (0.5x FN): Test MR | Better safe than sorry (0.5x FP): Training | Better safe than sorry (0.5x FP): Training MR | Better safe than sorry (0.5x FP): Test | Better safe than sorry (0.5x FP): Test MR |
|---|---|---|---|---|---|---|---|---|---|---|---|---|
| Adaptive: CHOZ | 81 | 1 | 86 | 0 | 84 | 0 | 88 | 1 | 82 | 0 | 85 | 0 |
| Adaptive: CHO | 79 | 4 | 86 | 1 | 82 | 4 | 89 | 0 | 80 | 4 | 84 | 2 |
| Adaptive: HZ | 80 | 3 | 86 | 2 | 83 | 1 | 88 | 2 | 81 | 3 | 85 | 1 |
| Simplified adaptive: HZ | 79 | 3 | 86 | 1 | 83 | 3 | 89 | 0 | 79 | 4 | 84 | 1 |
| Community Levels | 80 | 3 | 75 | 23 | 82 | 3 | 72 | 34 | 79 | 9 | 77 | 13 |
| Z | 77 | 5 | 80 | 11 | 78 | 12 | 78 | 16 | 77 | 8 | 81 | 10 |
| CHO | 80 | 4 | 83 | 6 | 83 | 1 | 85 | 8 | 79 | 5 | 80 | 11 |
| HO | 80 | 4 | 83 | 6 | 83 | 2 | 88 | 1 | 79 | 5 | 80 | 12 |
| CH | 80 | 5 | 82 | 8 | 83 | 1 | 84 | 11 | 80 | 4 | 74 | 22 |
| H | 80 | 4 | 82 | 9 | 82 | 3 | 87 | 2 | 79 | 4 | 74 | 23 |
| C | 80 | 4 | 65 | 32 | 83 | 3 | 71 | 27 | 79 | 5 | 66 | 28 |
| Prevalence | 35 | 71 | 26 | 82 | 35 | 76 | 26 | 85 | 35 | 69 | 26 | 79 |

**Fig. S4.** Head-to-head comparison results. The top plots display results from state-level analyses and the bottom plots display results from county-level analyses, both weighted for population. Metrics are displayed on the left, with training data from Q2-Q4 2021 and test data from Q1-Q3 2022. Cells report weighted accuracy and maximum regret (MR) over training and test periods. Rows vary outcomes, and columns vary preferences for false positive versus false negatives, with "neutral" corresponding to unweighted accuracy. Prevalence indicates the proportion of high location-weeks in a given time period. Weighted accuracy by quarter, including for intensive care usage, is presented in Figures S5-S7.

States

**>1 death/100K/wk — Neutral**

| | 21-2 | 21-3 | 21-4 | 22-1 | 22-2 | 22-3 | Test | Training | Overall |
|---|---|---|---|---|---|---|---|---|---|
| Adaptive: CHOZ | 81 | 90 | 92 | 85 | 84 | 80 | 83 | 88 | 85 |
| Adaptive: CHO | 80 | 90 | 93 | 85 | 83 | 73 | 80 | 88 | 84 |
| Adaptive: HZ | 80 | 92 | 94 | 85 | 84 | 81 | 83 | 89 | 86 |
| Simplified adaptive: HZ | 75 | 89 | 94 | 82 | 81 | 83 | 82 | 86 | 84 |
| Community Levels | 71 | 72 | 50 | 75 | 79 | 59 | 71 | 64 | 68 |
| Z | 72 | 78 | 91 | 82 | 81 | 80 | 81 | 80 | 81 |
| CHO | 79 | 90 | 87 | 87 | 75 | 42 | 68 | 86 | 77 |
| HO | 79 | 90 | 87 | 87 | 75 | 42 | 68 | 85 | 77 |
| CH | 76 | 93 | 93 | 88 | 48 | 31 | 56 | 87 | 72 |
| H | 66 | 91 | 94 | 90 | 49 | 31 | 56 | 83 | 70 |
| C | 76 | 89 | 93 | 80 | 24 | 29 | 45 | 86 | 65 |
| Prevalence | 35 | 75 | 94 | 78 | 17 | 29 | 41 | 68 | 55 |

**>1 death/100K/wk — Don't cry wolf (0.5x FN)**

| | 21-2 | 21-3 | 21-4 | 22-1 | 22-2 | 22-3 | Test | Training | Overall |
|---|---|---|---|---|---|---|---|---|---|
| Adaptive: CHOZ | 83 | 88 | 91 | 87 | 89 | 84 | 87 | 88 | 87 |
| Adaptive: CHO | 81 | 88 | 92 | 87 | 89 | 83 | 86 | 87 | 87 |
| Adaptive: HZ | 82 | 93 | 92 | 88 | 90 | 84 | 87 | 89 | 88 |
| Simplified adaptive: HZ | 73 | 89 | 93 | 84 | 85 | 84 | 84 | 85 | 85 |
| Community Levels | 81 | 81 | 66 | 83 | 80 | 55 | 72 | 76 | 74 |
| Z | 66 | 85 | 91 | 77 | 81 | 80 | 79 | 81 | 80 |
| CHO | 80 | 92 | 90 | 89 | 75 | 26 | 63 | 87 | 75 |
| HO | 79 | 92 | 90 | 89 | 75 | 26 | 63 | 87 | 75 |
| CH | 81 | 92 | 85 | 86 | 36 | 17 | 47 | 86 | 66 |
| H | 85 | 93 | 75 | 87 | 79 | 39 | 68 | 84 | 76 |
| C | 81 | 92 | 85 | 84 | 23 | 17 | 41 | 86 | 64 |
| Prevalence | 35 | 75 | 94 | 78 | 17 | 29 | 41 | 68 | 55 |

**>1 death/100K/wk — Better safe than sorry (0.5x FP)**

| | 21-2 | 21-3 | 21-4 | 22-1 | 22-2 | 22-3 | Test | Training | Overall |
|---|---|---|---|---|---|---|---|---|---|
| Adaptive: CHOZ | 83 | 93 | 94 | 80 | 82 | 76 | 80 | 90 | 85 |
| Adaptive: CHO | 81 | 93 | 96 | 81 | 78 | 66 | 75 | 90 | 82 |
| Adaptive: HZ | 84 | 92 | 96 | 82 | 82 | 80 | 81 | 91 | 86 |
| Simplified adaptive: HZ | 79 | 91 | 96 | 86 | 79 | 80 | 82 | 89 | 85 |
| Community Levels | 62 | 62 | 34 | 66 | 79 | 64 | 70 | 53 | 61 |
| Z | 78 | 70 | 91 | 87 | 81 | 80 | 83 | 80 | 81 |
| CHO | 79 | 88 | 85 | 85 | 76 | 58 | 73 | 84 | 78 |
| HO | 79 | 88 | 85 | 85 | 76 | 58 | 73 | 84 | 78 |
| CH | 81 | 95 | 94 | 88 | 63 | 54 | 68 | 90 | 79 |
| H | 76 | 94 | 96 | 90 | 63 | 54 | 69 | 88 | 79 |
| C | 82 | 93 | 94 | 84 | 49 | 53 | 62 | 90 | 76 |
| Prevalence | 35 | 75 | 94 | 78 | 17 | 29 | 41 | 68 | 55 |

**>2 deaths/100K/wk — Neutral**

| | 21-2 | 21-3 | 21-4 | 22-1 | 22-2 | 22-3 | Test | Training | Overall |
|---|---|---|---|---|---|---|---|---|---|
| Adaptive: CHOZ | 96 | 86 | 82 | 87 | 96 | 93 | 92 | 88 | 90 |
| Adaptive: CHO | 95 | 84 | 81 | 87 | 96 | 96 | 93 | 87 | 90 |
| Adaptive: HZ | 95 | 87 | 88 | 87 | 97 | 95 | 93 | 90 | 91 |
| Simplified adaptive: HZ | 93 | 83 | 86 | 90 | 97 | 96 | 94 | 87 | 91 |
| Community Levels | 96 | 88 | 81 | 87 | 85 | 60 | 77 | 88 | 83 |
| Z | 92 | 81 | 77 | 71 | 95 | 96 | 87 | 83 | 85 |
| CHO | 96 | 89 | 81 | 86 | 84 | 59 | 76 | 89 | 83 |
| HO | 95 | 88 | 82 | 88 | 96 | 91 | 91 | 88 | 90 |
| CH | 96 | 89 | 80 | 86 | 66 | 57 | 70 | 88 | 79 |
| H | 94 | 88 | 82 | 88 | 95 | 90 | 91 | 88 | 90 |
| C | 96 | 89 | 80 | 86 | 66 | 57 | 70 | 88 | 79 |
| Prevalence | 5 | 47 | 56 | 61 | 3 | 5 | 23 | 36 | 29 |

**>2 deaths/100K/wk — Don't cry wolf (0.5x FN)**

| | 21-2 | 21-3 | 21-4 | 22-1 | 22-2 | 22-3 | Test | Training | Overall |
|---|---|---|---|---|---|---|---|---|---|
| Adaptive: CHOZ | 96 | 86 | 83 | 88 | 97 | 95 | 93 | 88 | 91 |
| Adaptive: CHO | 95 | 84 | 83 | 88 | 97 | 98 | 94 | 87 | 91 |
| Adaptive: HZ | 96 | 89 | 88 | 86 | 97 | 95 | 93 | 91 | 92 |
| Simplified adaptive: HZ | 94 | 87 | 87 | 89 | 98 | 96 | 94 | 89 | 92 |
| Community Levels | 96 | 89 | 84 | 89 | 80 | 47 | 72 | 90 | 81 |
| Z | 91 | 86 | 78 | 66 | 95 | 96 | 86 | 85 | 85 |
| CHO | 96 | 90 | 84 | 89 | 87 | 49 | 75 | 90 | 83 |
| HO | 94 | 89 | 87 | 90 | 95 | 88 | 91 | 90 | 91 |
| CH | 95 | 89 | 86 | 90 | 95 | 88 | 91 | 90 | 90 |
| H | 92 | 89 | 87 | 90 | 95 | 88 | 91 | 89 | 90 |
| C | 96 | 89 | 84 | 88 | 55 | 43 | 62 | 90 | 76 |
| Prevalence | 5 | 47 | 56 | 61 | 3 | 5 | 23 | 36 | 29 |

**>2 deaths/100K/wk — Better safe than sorry (0.5x FP)**

| | 21-2 | 21-3 | 21-4 | 22-1 | 22-2 | 22-3 | Test | Training | Overall |
|---|---|---|---|---|---|---|---|---|---|
| Adaptive: CHOZ | 95 | 88 | 81 | 87 | 95 | 92 | 92 | 88 | 90 |
| Adaptive: CHO | 95 | 86 | 81 | 87 | 95 | 95 | 93 | 87 | 90 |
| Adaptive: HZ | 95 | 87 | 87 | 88 | 95 | 94 | 92 | 90 | 91 |
| Simplified adaptive: HZ | 94 | 81 | 88 | 89 | 95 | 96 | 93 | 88 | 91 |
| Community Levels | 97 | 88 | 77 | 84 | 89 | 73 | 82 | 87 | 85 |
| Z | 94 | 75 | 77 | 75 | 95 | 97 | 89 | 82 | 86 |
| CHO | 94 | 88 | 83 | 86 | 87 | 59 | 77 | 88 | 83 |
| HO | 89 | 87 | 89 | 90 | 92 | 62 | 81 | 88 | 85 |
| CH | 94 | 87 | 82 | 86 | 88 | 62 | 79 | 88 | 83 |
| H | 88 | 86 | 88 | 90 | 88 | 59 | 79 | 87 | 83 |
| C | 94 | 88 | 82 | 86 | 64 | 55 | 68 | 88 | 78 |
| Prevalence | 5 | 47 | 56 | 61 | 3 | 5 | 23 | 36 | 29 |

**>2 ICU patients/100K/wk — Neutral**

| | 21-2 | 21-3 | 21-4 | 22-1 | 22-2 | 22-3 | Test | Training | Overall |
|---|---|---|---|---|---|---|---|---|---|
| Adaptive: CHOZ | 75 | 92 | 90 | 86 | 92 | 94 | 91 | 86 | 88 |
| Adaptive: CHO | 78 | 92 | 90 | 90 | 94 | 94 | 93 | 87 | 90 |
| Adaptive: HZ | 76 | 88 | 95 | 86 | 98 | 94 | 93 | 86 | 89 |
| Simplified adaptive: HZ | 80 | 81 | 92 | 74 | 99 | 94 | 89 | 85 | 87 |
| Community Levels | 68 | 58 | 53 | 91 | 86 | 61 | 79 | 60 | 70 |
| Z | 76 | 83 | 93 | 77 | 99 | 94 | 90 | 84 | 87 |
| CHO | 81 | 79 | 91 | 73 | 82 | 27 | 61 | 84 | 72 |
| HO | 80 | 79 | 91 | 73 | 82 | 27 | 61 | 83 | 72 |
| CH | 77 | 92 | 95 | 67 | 41 | 7 | 38 | 88 | 63 |
| H | 67 | 93 | 93 | 65 | 40 | 6 | 37 | 84 | 61 |
| C | 75 | 93 | 94 | 55 | 11 | 5 | 24 | 87 | 56 |
| Prevalence | 37 | 89 | 92 | 45 | 1 | 5 | 17 | 73 | 45 |

**>2 ICU patients/100K/wk — Don't cry wolf (0.5x FN)**

| | 21-2 | 21-3 | 21-4 | 22-1 | 22-2 | 22-3 | Test | Training | Overall |
|---|---|---|---|---|---|---|---|---|---|
| Adaptive: CHOZ | 77 | 92 | 92 | 82 | 91 | 95 | 89 | 87 | 88 |
| Adaptive: CHO | 81 | 92 | 91 | 87 | 94 | 96 | 92 | 88 | 90 |
| Adaptive: HZ | 77 | 90 | 92 | 82 | 98 | 95 | 92 | 87 | 89 |
| Simplified adaptive: HZ | 84 | 84 | 90 | 68 | 99 | 95 | 87 | 86 | 87 |
| Community Levels | 79 | 72 | 69 | 88 | 81 | 48 | 73 | 73 | 73 |
| Z | 73 | 88 | 93 | 69 | 99 | 93 | 87 | 85 | 86 |
| CHO | 82 | 86 | 93 | 64 | 76 | 3 | 48 | 87 | 67 |
| HO | 81 | 86 | 93 | 64 | 76 | 3 | 48 | 87 | 67 |
| CH | 83 | 89 | 88 | 72 | 23 | 0 | 27 | 87 | 57 |
| H | 83 | 85 | 76 | 77 | 80 | 19 | 59 | 81 | 70 |
| C | 83 | 89 | 88 | 68 | 10 | 0 | 22 | 87 | 54 |
| Prevalence | 37 | 89 | 92 | 45 | 1 | 5 | 17 | 73 | 45 |

**>2 ICU patients/100K/wk — Better safe than sorry (0.5x FP)**

| | 21-2 | 21-3 | 21-4 | 22-1 | 22-2 | 22-3 | Test | Training | Overall |
|---|---|---|---|---|---|---|---|---|---|
| Adaptive: CHOZ | 78 | 92 | 89 | 90 | 94 | 94 | 93 | 86 | 90 |
| Adaptive: CHO | 79 | 92 | 89 | 93 | 95 | 94 | 94 | 86 | 90 |
| Adaptive: HZ | 79 | 88 | 96 | 90 | 97 | 94 | 94 | 87 | 91 |
| Simplified adaptive: HZ | 80 | 80 | 95 | 80 | 98 | 94 | 91 | 85 | 88 |
| Community Levels | 58 | 44 | 38 | 94 | 90 | 74 | 86 | 47 | 66 |
| Z | 80 | 77 | 93 | 85 | 98 | 94 | 92 | 83 | 88 |
| CHO | 79 | 72 | 89 | 82 | 88 | 51 | 74 | 80 | 77 |
| HO | 79 | 72 | 89 | 82 | 88 | 51 | 74 | 80 | 77 |
| CH | 81 | 89 | 96 | 78 | 60 | 38 | 59 | 89 | 74 |
| H | 76 | 91 | 95 | 77 | 60 | 38 | 58 | 88 | 73 |
| C | 80 | 92 | 96 | 70 | 41 | 37 | 49 | 89 | 69 |
| Prevalence | 37 | 89 | 92 | 45 | 1 | 5 | 17 | 73 | 45 |

**Fig. S5.** State-level results by quarter. Metrics are displayed on the left, with training data from Q3-Q4 2021 and test data from Q1-Q3 2022. Cells report weighted accuracy. Preferences for false positive versus false negatives varying across columns (with "neutral" corresponding to unweighted accuracy) and outcomes across rows. Prevalence indicates the proportion of high location weeks in a given quarter.

HSAs

**>1 death/100K/wk — Neutral**

| | 21-2 | 21-3 | 21-4 | 22-1 | 22-2 | 22-3 | Test | Training | Overall |
|---|---|---|---|---|---|---|---|---|---|
| Adaptive: CHOZ | 76 | 87 | 86 | 84 | 74 | 71 | 77 | 83 | 80 |
| Adaptive: CHO | 72 | 86 | 84 | 82 | 73 | 66 | 73 | 80 | 77 |
| Adaptive: HZ | 76 | 84 | 87 | 84 | 74 | 72 | 77 | 82 | 80 |
| Simplified adaptive: HZ | 73 | 84 | 86 | 84 | 73 | 72 | 76 | 81 | 78 |
| Community Levels | 63 | 72 | 56 | 71 | 71 | 62 | 68 | 64 | 66 |
| Z | 72 | 78 | 83 | 82 | 72 | 72 | 75 | 78 | 77 |
| CHO | 70 | 82 | 72 | 78 | 72 | 59 | 70 | 75 | 72 |
| HO | 70 | 82 | 72 | 78 | 72 | 59 | 70 | 75 | 72 |
| CH | 73 | 85 | 83 | 82 | 49 | 41 | 58 | 81 | 69 |
| H | 70 | 84 | 84 | 84 | 49 | 41 | 58 | 80 | 69 |
| C | 72 | 82 | 84 | 78 | 32 | 39 | 50 | 79 | 64 |
| Prevalence | 44 | 70 | 84 | 79 | 27 | 39 | 48 | 66 | 57 |

**>1 death/100K/wk — Don't cry wolf (0.5x FN)**

| | 21-2 | 21-3 | 21-4 | 22-1 | 22-2 | 22-3 | Test | Training | Overall |
|---|---|---|---|---|---|---|---|---|---|
| Adaptive: CHOZ | 79 | 87 | 83 | 83 | 82 | 77 | 81 | 83 | 82 |
| Adaptive: CHO | 76 | 84 | 83 | 84 | 82 | 75 | 80 | 81 | 81 |
| Adaptive: HZ | 79 | 86 | 84 | 83 | 83 | 77 | 81 | 83 | 82 |
| Simplified adaptive: HZ | 72 | 85 | 83 | 84 | 80 | 77 | 80 | 80 | 80 |
| Community Levels | 75 | 81 | 70 | 80 | 75 | 64 | 73 | 75 | 74 |
| Z | 68 | 84 | 83 | 79 | 71 | 72 | 74 | 78 | 76 |
| CHO | 78 | 85 | 80 | 83 | 78 | 59 | 73 | 81 | 77 |
| HO | 78 | 85 | 80 | 83 | 78 | 59 | 73 | 81 | 77 |
| CH | 77 | 85 | 81 | 83 | 43 | 30 | 52 | 81 | 66 |
| H | 79 | 85 | 78 | 82 | 72 | 51 | 68 | 81 | 74 |
| C | 76 | 84 | 81 | 82 | 29 | 28 | 46 | 80 | 63 |
| Prevalence | 44 | 70 | 84 | 79 | 27 | 39 | 48 | 66 | 57 |

**>1 death/100K/wk — Better safe than sorry (0.5x FP)**

| | 21-2 | 21-3 | 21-4 | 22-1 | 22-2 | 22-3 | Test | Training | Overall |
|---|---|---|---|---|---|---|---|---|---|
| Adaptive: CHOZ | 79 | 89 | 90 | 89 | 74 | 72 | 78 | 86 | 82 |
| Adaptive: CHO | 74 | 87 | 89 | 86 | 63 | 62 | 70 | 84 | 77 |
| Adaptive: HZ | 78 | 87 | 91 | 89 | 74 | 72 | 78 | 86 | 82 |
| Simplified adaptive: HZ | 78 | 86 | 90 | 88 | 74 | 72 | 78 | 85 | 81 |
| Community Levels | 51 | 64 | 42 | 62 | 67 | 61 | 63 | 52 | 58 |
| Z | 76 | 73 | 83 | 86 | 73 | 71 | 77 | 77 | 77 |
| CHO | 62 | 78 | 65 | 73 | 67 | 60 | 66 | 68 | 67 |
| HO | 62 | 78 | 65 | 73 | 67 | 60 | 66 | 68 | 67 |
| CH | 73 | 88 | 83 | 81 | 59 | 59 | 66 | 81 | 74 |
| H | 74 | 88 | 85 | 83 | 59 | 59 | 67 | 82 | 74 |
| C | 74 | 87 | 88 | 81 | 51 | 59 | 64 | 83 | 73 |
| Prevalence | 44 | 70 | 84 | 79 | 27 | 39 | 48 | 66 | 57 |

**>2 deaths/100K/wk — Neutral**

| | 21-2 | 21-3 | 21-4 | 22-1 | 22-2 | 22-3 | Test | Training | Overall |
|---|---|---|---|---|---|---|---|---|---|
| Adaptive: CHOZ | 89 | 84 | 79 | 82 | 93 | 90 | 88 | 84 | 86 |
| Adaptive: CHO | 88 | 82 | 76 | 82 | 93 | 90 | 88 | 82 | 85 |
| Adaptive: HZ | 89 | 81 | 80 | 81 | 93 | 90 | 88 | 83 | 86 |
| Simplified adaptive: HZ | 88 | 80 | 77 | 80 | 92 | 90 | 87 | 82 | 85 |
| Community Levels | 89 | 85 | 75 | 81 | 80 | 66 | 76 | 83 | 79 |
| Z | 83 | 80 | 76 | 71 | 88 | 88 | 82 | 80 | 81 |
| CHO | 87 | 83 | 76 | 80 | 89 | 71 | 80 | 82 | 81 |
| HO | 89 | 83 | 73 | 78 | 91 | 85 | 85 | 82 | 83 |
| CH | 87 | 83 | 77 | 81 | 78 | 56 | 72 | 82 | 77 |
| H | 89 | 83 | 73 | 78 | 90 | 81 | 83 | 82 | 82 |
| C | 88 | 84 | 73 | 80 | 62 | 57 | 66 | 82 | 74 |
| Prevalence | 13 | 42 | 56 | 62 | 7 | 10 | 26 | 37 | 32 |

**>2 deaths/100K/wk — Don't cry wolf (0.5x FN)**

| | 21-2 | 21-3 | 21-4 | 22-1 | 22-2 | 22-3 | Test | Training | Overall |
|---|---|---|---|---|---|---|---|---|---|
| Adaptive: CHOZ | 92 | 84 | 82 | 83 | 95 | 94 | 91 | 86 | 88 |
| Adaptive: CHO | 92 | 81 | 79 | 83 | 95 | 93 | 91 | 84 | 87 |
| Adaptive: HZ | 92 | 83 | 81 | 80 | 95 | 94 | 90 | 85 | 88 |
| Simplified adaptive: HZ | 92 | 81 | 77 | 83 | 95 | 93 | 91 | 83 | 87 |
| Community Levels | 90 | 84 | 80 | 84 | 77 | 57 | 73 | 85 | 79 |
| Z | 81 | 85 | 77 | 67 | 88 | 88 | 81 | 81 | 81 |
| CHO | 91 | 85 | 79 | 83 | 93 | 84 | 86 | 85 | 86 |
| HO | 91 | 84 | 80 | 83 | 93 | 84 | 86 | 85 | 86 |
| CH | 92 | 85 | 78 | 83 | 92 | 82 | 85 | 85 | 85 |
| H | 90 | 84 | 80 | 83 | 91 | 79 | 84 | 84 | 84 |
| C | 92 | 86 | 77 | 82 | 69 | 67 | 73 | 85 | 79 |
| Prevalence | 13 | 42 | 56 | 62 | 7 | 10 | 26 | 37 | 32 |

**>2 deaths/100K/wk — Better safe than sorry (0.5x FP)**

| | 21-2 | 21-3 | 21-4 | 22-1 | 22-2 | 22-3 | Test | Training | Overall |
|---|---|---|---|---|---|---|---|---|---|
| Adaptive: CHOZ | 87 | 85 | 82 | 84 | 91 | 88 | 88 | 85 | 86 |
| Adaptive: CHO | 84 | 85 | 78 | 82 | 91 | 88 | 87 | 83 | 85 |
| Adaptive: HZ | 87 | 81 | 83 | 83 | 91 | 88 | 87 | 84 | 85 |
| Simplified adaptive: HZ | 86 | 81 | 79 | 83 | 89 | 88 | 86 | 82 | 84 |
| Community Levels | 87 | 85 | 70 | 78 | 83 | 74 | 78 | 81 | 79 |
| Z | 85 | 76 | 76 | 75 | 89 | 88 | 84 | 79 | 82 |
| CHO | 85 | 83 | 78 | 81 | 87 | 71 | 80 | 82 | 81 |
| HO | 85 | 84 | 76 | 80 | 88 | 76 | 81 | 82 | 81 |
| CH | 85 | 84 | 78 | 80 | 81 | 65 | 75 | 82 | 79 |
| H | 83 | 83 | 78 | 80 | 81 | 64 | 75 | 82 | 78 |
| C | 85 | 84 | 75 | 80 | 62 | 57 | 66 | 82 | 74 |
| Prevalence | 13 | 42 | 56 | 62 | 7 | 10 | 26 | 37 | 32 |

**>2 ICU patients/100K/wk — Neutral**

| | 21-2 | 21-3 | 21-4 | 22-1 | 22-2 | 22-3 | Test | Training | Overall |
|---|---|---|---|---|---|---|---|---|---|
| Adaptive: CHOZ | 78 | 84 | 84 | 84 | 96 | 93 | 91 | 82 | 87 |
| Adaptive: CHO | 79 | 83 | 81 | 84 | 96 | 92 | 91 | 81 | 86 |
| Adaptive: HZ | 78 | 80 | 83 | 82 | 97 | 93 | 91 | 80 | 85 |
| Simplified adaptive: HZ | 79 | 79 | 83 | 74 | 97 | 92 | 88 | 80 | 84 |
| Community Levels | 74 | 61 | 61 | 85 | 85 | 68 | 80 | 66 | 73 |
| Z | 79 | 80 | 82 | 76 | 97 | 92 | 89 | 80 | 84 |
| CHO | 77 | 72 | 77 | 76 | 89 | 61 | 75 | 75 | 75 |
| HO | 77 | 72 | 77 | 76 | 89 | 61 | 75 | 75 | 75 |
| CH | 74 | 84 | 85 | 67 | 47 | 15 | 43 | 81 | 62 |
| H | 69 | 86 | 85 | 64 | 46 | 15 | 42 | 80 | 61 |
| C | 77 | 79 | 83 | 71 | 37 | 19 | 42 | 80 | 61 |
| Prevalence | 31 | 81 | 76 | 40 | 3 | 8 | 17 | 63 | 40 |

**>2 ICU patients/100K/wk — Don't cry wolf (0.5x FN)**

| | 21-2 | 21-3 | 21-4 | 22-1 | 22-2 | 22-3 | Test | Training | Overall |
|---|---|---|---|---|---|---|---|---|---|
| Adaptive: CHOZ | 81 | 84 | 84 | 81 | 97 | 94 | 91 | 83 | 87 |
| Adaptive: CHO | 83 | 83 | 81 | 80 | 97 | 95 | 90 | 82 | 86 |
| Adaptive: HZ | 80 | 82 | 84 | 80 | 98 | 94 | 90 | 82 | 86 |
| Simplified adaptive: HZ | 82 | 75 | 82 | 68 | 98 | 93 | 86 | 80 | 83 |
| Community Levels | 82 | 73 | 73 | 82 | 81 | 59 | 74 | 76 | 75 |
| Z | 76 | 84 | 83 | 69 | 97 | 92 | 86 | 81 | 84 |
| CHO | 82 | 79 | 82 | 72 | 87 | 52 | 70 | 81 | 76 |
| HO | 81 | 79 | 82 | 68 | 87 | 50 | 68 | 81 | 75 |
| CH | 81 | 82 | 85 | 66 | 35 | 0 | 33 | 83 | 58 |
| H | 82 | 80 | 79 | 75 | 76 | 36 | 62 | 80 | 71 |
| C | 80 | 82 | 84 | 62 | 16 | 0 | 23 | 82 | 52 |
| Prevalence | 31 | 81 | 76 | 40 | 3 | 8 | 17 | 63 | 40 |

**>2 ICU patients/100K/wk — Better safe than sorry (0.5x FP)**

| | 21-2 | 21-3 | 21-4 | 22-1 | 22-2 | 22-3 | Test | Training | Overall |
|---|---|---|---|---|---|---|---|---|---|
| Adaptive: CHOZ | 80 | 85 | 85 | 88 | 96 | 92 | 92 | 83 | 88 |
| Adaptive: CHO | 78 | 85 | 87 | 88 | 95 | 90 | 91 | 83 | 87 |
| Adaptive: HZ | 80 | 81 | 84 | 86 | 96 | 92 | 91 | 82 | 87 |
| Simplified adaptive: HZ | 81 | 75 | 84 | 75 | 96 | 92 | 88 | 80 | 84 |
| Community Levels | 67 | 49 | 50 | 89 | 89 | 77 | 85 | 55 | 70 |
| Z | 81 | 75 | 81 | 84 | 96 | 93 | 91 | 79 | 85 |
| CHO | 73 | 64 | 72 | 84 | 91 | 71 | 82 | 70 | 76 |
| HO | 73 | 64 | 72 | 83 | 91 | 71 | 82 | 70 | 76 |
| CH | 79 | 83 | 88 | 77 | 65 | 43 | 62 | 83 | 72 |
| H | 77 | 85 | 88 | 76 | 64 | 43 | 61 | 83 | 72 |
| C | 76 | 86 | 86 | 68 | 46 | 40 | 51 | 83 | 67 |
| Prevalence | 31 | 81 | 76 | 40 | 3 | 8 | 17 | 63 | 40 |

**Fig. S6.** HSA-level results by quarter. Metrics are displayed on the left, with training data from Q3-Q4 2021 and test data from Q1-Q3 2022. Cells report weighted accuracy. Preferences for false positive versus false negatives varying across columns (with "neutral" corresponding to unweighted accuracy) and outcomes across rows. Prevalence indicates the proportion of high location weeks in a given quarter.

Counties

**Neutral — >1 death/100K/wk**

| | 21-2 | 21-3 | 21-4 | 22-1 | 22-2 | 22-3 | Test | Training | Overall |
|---|---|---|---|---|---|---|---|---|---|
| Adaptive: CHOZ | 73 | 81 | 79 | 81 | 76 | 69 | 75 | 77 | 76 |
| Adaptive: CHO | 71 | 79 | 77 | 78 | 75 | 67 | 73 | 76 | 75 |
| Adaptive: HZ | 72 | 77 | 79 | 80 | 75 | 69 | 75 | 76 | 75 |
| Simplified adaptive: HZ | 69 | 77 | 77 | 80 | 72 | 69 | 74 | 75 | 74 |
| Community Levels | 68 | 72 | 60 | 72 | 72 | 62 | 69 | 67 | 68 |
| Z | 69 | 75 | 76 | 77 | 72 | 67 | 72 | 73 | 73 |
| CHO | 72 | 77 | 72 | 78 | 73 | 60 | 70 | 74 | 72 |
| HO | 72 | 77 | 72 | 78 | 73 | 60 | 70 | 74 | 72 |
| CH | 70 | 78 | 78 | 80 | 51 | 38 | 57 | 75 | 66 |
| H | 65 | 77 | 78 | 81 | 50 | 38 | 56 | 74 | 65 |
| C | 71 | 79 | 75 | 77 | 43 | 42 | 54 | 75 | 64 |
| Prevalence | 37 | 63 | 75 | 74 | 25 | 35 | 45 | 59 | 52 |

**Don't cry wolf (0.5x FN) — >1 death/100K/wk**

| | 21-2 | 21-3 | 21-4 | 22-1 | 22-2 | 22-3 | Test | Training | Overall |
|---|---|---|---|---|---|---|---|---|---|
| Adaptive: CHOZ | 79 | 82 | 78 | 80 | 83 | 78 | 80 | 79 | 80 |
| Adaptive: CHO | 78 | 79 | 78 | 79 | 83 | 77 | 80 | 78 | 79 |
| Adaptive: HZ | 78 | 80 | 77 | 79 | 83 | 78 | 80 | 78 | 79 |
| Simplified adaptive: HZ | 75 | 77 | 76 | 81 | 80 | 78 | 80 | 76 | 78 |
| Community Levels | 78 | 78 | 71 | 79 | 76 | 63 | 73 | 76 | 74 |
| Z | 65 | 80 | 76 | 74 | 71 | 67 | 71 | 74 | 72 |
| CHO | 78 | 79 | 77 | 81 | 79 | 59 | 73 | 78 | 75 |
| HO | 77 | 79 | 77 | 81 | 79 | 59 | 73 | 78 | 75 |
| CH | 78 | 80 | 76 | 81 | 73 | 51 | 68 | 78 | 73 |
| H | 76 | 79 | 76 | 81 | 72 | 49 | 67 | 77 | 72 |
| C | 78 | 80 | 74 | 79 | 44 | 39 | 54 | 77 | 66 |
| Prevalence | 37 | 63 | 75 | 74 | 25 | 35 | 45 | 59 | 52 |

**Better safe than sorry (0.5x FP) — >1 death/100K/wk**

| | 21-2 | 21-3 | 21-4 | 22-1 | 22-2 | 22-3 | Test | Training | Overall |
|---|---|---|---|---|---|---|---|---|---|
| Adaptive: CHOZ | 75 | 84 | 84 | 86 | 73 | 67 | 75 | 81 | 78 |
| Adaptive: CHO | 71 | 83 | 84 | 83 | 66 | 61 | 70 | 79 | 75 |
| Adaptive: HZ | 74 | 79 | 84 | 86 | 73 | 67 | 75 | 79 | 77 |
| Simplified adaptive: HZ | 73 | 80 | 84 | 85 | 72 | 67 | 75 | 79 | 77 |
| Community Levels | 59 | 66 | 48 | 65 | 68 | 62 | 65 | 57 | 61 |
| Z | 72 | 70 | 76 | 80 | 72 | 67 | 73 | 73 | 73 |
| CHO | 66 | 76 | 67 | 74 | 68 | 62 | 68 | 70 | 69 |
| HO | 66 | 76 | 67 | 74 | 68 | 62 | 68 | 70 | 69 |
| CH | 72 | 83 | 80 | 80 | 61 | 57 | 66 | 78 | 72 |
| H | 71 | 83 | 82 | 82 | 61 | 57 | 66 | 79 | 72 |
| C | 72 | 82 | 82 | 79 | 53 | 57 | 63 | 79 | 71 |
| Prevalence | 37 | 63 | 75 | 74 | 25 | 35 | 45 | 59 | 52 |

**Neutral — >2 deaths/100K/wk**

| | 21-2 | 21-3 | 21-4 | 22-1 | 22-2 | 22-3 | Test | Training | Overall |
|---|---|---|---|---|---|---|---|---|---|
| Adaptive: CHOZ | 86 | 81 | 76 | 79 | 91 | 89 | 86 | 81 | 84 |
| Adaptive: CHO | 85 | 79 | 72 | 78 | 91 | 88 | 86 | 79 | 82 |
| Adaptive: HZ | 86 | 79 | 75 | 77 | 91 | 89 | 86 | 80 | 83 |
| Simplified adaptive: HZ | 85 | 79 | 73 | 78 | 91 | 88 | 86 | 79 | 82 |
| Community Levels | 86 | 82 | 73 | 78 | 80 | 66 | 75 | 80 | 78 |
| Z | 81 | 79 | 73 | 68 | 86 | 85 | 80 | 77 | 79 |
| CHO | 86 | 81 | 72 | 77 | 90 | 83 | 83 | 80 | 82 |
| HO | 86 | 81 | 72 | 77 | 90 | 83 | 83 | 80 | 82 |
| CH | 86 | 82 | 71 | 76 | 89 | 81 | 82 | 80 | 81 |
| H | 86 | 81 | 72 | 77 | 89 | 80 | 82 | 80 | 81 |
| C | 86 | 81 | 72 | 77 | 62 | 57 | 65 | 80 | 72 |
| Prevalence | 15 | 40 | 50 | 57 | 9 | 12 | 26 | 35 | 30 |

**Don't cry wolf (0.5x FN) — >2 deaths/100K/wk**

| | 21-2 | 21-3 | 21-4 | 22-1 | 22-2 | 22-3 | Test | Training | Overall |
|---|---|---|---|---|---|---|---|---|---|
| Adaptive: CHOZ | 91 | 83 | 78 | 79 | 94 | 92 | 88 | 84 | 86 |
| Adaptive: CHO | 90 | 79 | 76 | 80 | 94 | 92 | 89 | 82 | 85 |
| Adaptive: HZ | 91 | 82 | 78 | 78 | 94 | 92 | 88 | 83 | 86 |
| Simplified adaptive: HZ | 91 | 80 | 77 | 80 | 94 | 92 | 89 | 83 | 86 |
| Community Levels | 88 | 81 | 77 | 80 | 78 | 58 | 72 | 82 | 77 |
| Z | 79 | 82 | 73 | 64 | 86 | 85 | 78 | 78 | 78 |
| CHO | 90 | 83 | 77 | 80 | 92 | 84 | 85 | 83 | 84 |
| HO | 90 | 83 | 76 | 80 | 94 | 91 | 88 | 83 | 86 |
| CH | 90 | 83 | 77 | 80 | 91 | 81 | 84 | 83 | 84 |
| H | 90 | 81 | 75 | 79 | 93 | 90 | 87 | 82 | 85 |
| C | 90 | 83 | 75 | 79 | 69 | 65 | 71 | 83 | 77 |
| Prevalence | 15 | 40 | 50 | 57 | 9 | 12 | 26 | 35 | 30 |

**Better safe than sorry (0.5x FP) — >2 deaths/100K/wk**

| | 21-2 | 21-3 | 21-4 | 22-1 | 22-2 | 22-3 | Test | Training | Overall |
|---|---|---|---|---|---|---|---|---|---|
| Adaptive: CHOZ | 84 | 83 | 79 | 81 | 88 | 86 | 85 | 82 | 84 |
| Adaptive: CHO | 81 | 83 | 75 | 79 | 88 | 85 | 84 | 80 | 82 |
| Adaptive: HZ | 84 | 80 | 79 | 80 | 88 | 86 | 85 | 81 | 83 |
| Simplified adaptive: HZ | 84 | 79 | 75 | 80 | 87 | 86 | 84 | 79 | 82 |
| Community Levels | 84 | 83 | 70 | 77 | 82 | 73 | 77 | 79 | 78 |
| Z | 83 | 75 | 72 | 71 | 87 | 85 | 81 | 77 | 79 |
| CHO | 83 | 81 | 74 | 78 | 86 | 75 | 80 | 79 | 79 |
| HO | 82 | 81 | 74 | 78 | 86 | 74 | 80 | 79 | 79 |
| CH | 82 | 81 | 75 | 78 | 80 | 64 | 74 | 80 | 77 |
| H | 80 | 81 | 76 | 79 | 80 | 63 | 74 | 79 | 77 |
| C | 82 | 81 | 74 | 77 | 62 | 58 | 66 | 79 | 72 |
| Prevalence | 15 | 40 | 50 | 57 | 9 | 12 | 26 | 35 | 30 |

**Neutral — >2 ICU patients/100K/wk**

| | 21-2 | 21-3 | 21-4 | 22-1 | 22-2 | 22-3 | Test | Training | Overall |
|---|---|---|---|---|---|---|---|---|---|
| Adaptive: CHOZ | 78 | 83 | 84 | 84 | 97 | 93 | 91 | 81 | 86 |
| Adaptive: CHO | 79 | 82 | 81 | 84 | 96 | 92 | 91 | 81 | 86 |
| Adaptive: HZ | 78 | 80 | 83 | 82 | 97 | 93 | 91 | 80 | 85 |
| Simplified adaptive: HZ | 79 | 79 | 83 | 74 | 97 | 92 | 88 | 80 | 84 |
| Community Levels | 75 | 60 | 61 | 86 | 85 | 69 | 80 | 65 | 73 |
| Z | 79 | 80 | 82 | 76 | 97 | 92 | 89 | 80 | 84 |
| CHO | 77 | 72 | 77 | 76 | 89 | 61 | 75 | 75 | 75 |
| HO | 77 | 72 | 77 | 76 | 89 | 61 | 75 | 75 | 75 |
| CH | 74 | 84 | 85 | 67 | 48 | 16 | 44 | 81 | 62 |
| H | 69 | 86 | 85 | 64 | 46 | 15 | 42 | 80 | 61 |
| C | 77 | 78 | 82 | 71 | 37 | 20 | 43 | 79 | 61 |
| Prevalence | 31 | 81 | 76 | 40 | 3 | 8 | 17 | 63 | 40 |

**Don't cry wolf (0.5x FN) — >2 ICU patients/100K/wk**

| | 21-2 | 21-3 | 21-4 | 22-1 | 22-2 | 22-3 | Test | Training | Overall |
|---|---|---|---|---|---|---|---|---|---|
| Adaptive: CHOZ | 81 | 83 | 84 | 81 | 97 | 94 | 91 | 83 | 87 |
| Adaptive: CHO | 83 | 82 | 81 | 79 | 97 | 95 | 90 | 82 | 86 |
| Adaptive: HZ | 80 | 82 | 84 | 80 | 98 | 94 | 90 | 82 | 86 |
| Simplified adaptive: HZ | 82 | 75 | 82 | 68 | 98 | 93 | 86 | 80 | 83 |
| Community Levels | 82 | 73 | 73 | 82 | 82 | 61 | 75 | 76 | 75 |
| Z | 76 | 84 | 83 | 69 | 97 | 92 | 86 | 81 | 84 |
| CHO | 81 | 79 | 82 | 69 | 87 | 50 | 69 | 81 | 75 |
| HO | 81 | 79 | 82 | 68 | 87 | 50 | 68 | 81 | 75 |
| CH | 81 | 82 | 84 | 67 | 36 | 0 | 34 | 82 | 58 |
| H | 82 | 80 | 79 | 75 | 76 | 36 | 62 | 80 | 71 |
| C | 79 | 82 | 83 | 62 | 17 | 0 | 24 | 81 | 53 |
| Prevalence | 31 | 81 | 76 | 40 | 3 | 8 | 17 | 63 | 40 |

**Better safe than sorry (0.5x FP) — >2 ICU patients/100K/wk**

| | 21-2 | 21-3 | 21-4 | 22-1 | 22-2 | 22-3 | Test | Training | Overall |
|---|---|---|---|---|---|---|---|---|---|
| Adaptive: CHOZ | 80 | 84 | 85 | 88 | 96 | 92 | 92 | 83 | 87 |
| Adaptive: CHO | 77 | 84 | 86 | 88 | 95 | 90 | 91 | 82 | 87 |
| Adaptive: HZ | 80 | 81 | 84 | 86 | 96 | 92 | 91 | 82 | 86 |
| Simplified adaptive: HZ | 81 | 75 | 84 | 75 | 96 | 92 | 88 | 80 | 84 |
| Community Levels | 67 | 48 | 49 | 89 | 89 | 78 | 85 | 55 | 70 |
| Z | 81 | 75 | 81 | 84 | 96 | 93 | 91 | 79 | 85 |
| CHO | 73 | 64 | 72 | 84 | 91 | 71 | 82 | 70 | 76 |
| HO | 73 | 64 | 72 | 83 | 91 | 71 | 82 | 70 | 76 |
| CH | 78 | 83 | 87 | 78 | 65 | 44 | 62 | 82 | 72 |
| H | 77 | 85 | 88 | 76 | 64 | 43 | 61 | 83 | 72 |
| C | 75 | 86 | 85 | 69 | 47 | 40 | 52 | 82 | 67 |
| Prevalence | 31 | 81 | 76 | 40 | 3 | 8 | 17 | 63 | 40 |

**Fig. S7.** County-level results by quarter. Metrics are displayed on the left, with training data from Q3-Q4 2021 and test data from Q1-Q3 2022. Cells report weighted accuracy. Preferences for false positive versus false negatives varying across columns (with "neutral" corresponding to unweighted accuracy) and outcomes across rows. Prevalence indicates the proportion of high location weeks in a given quarter.

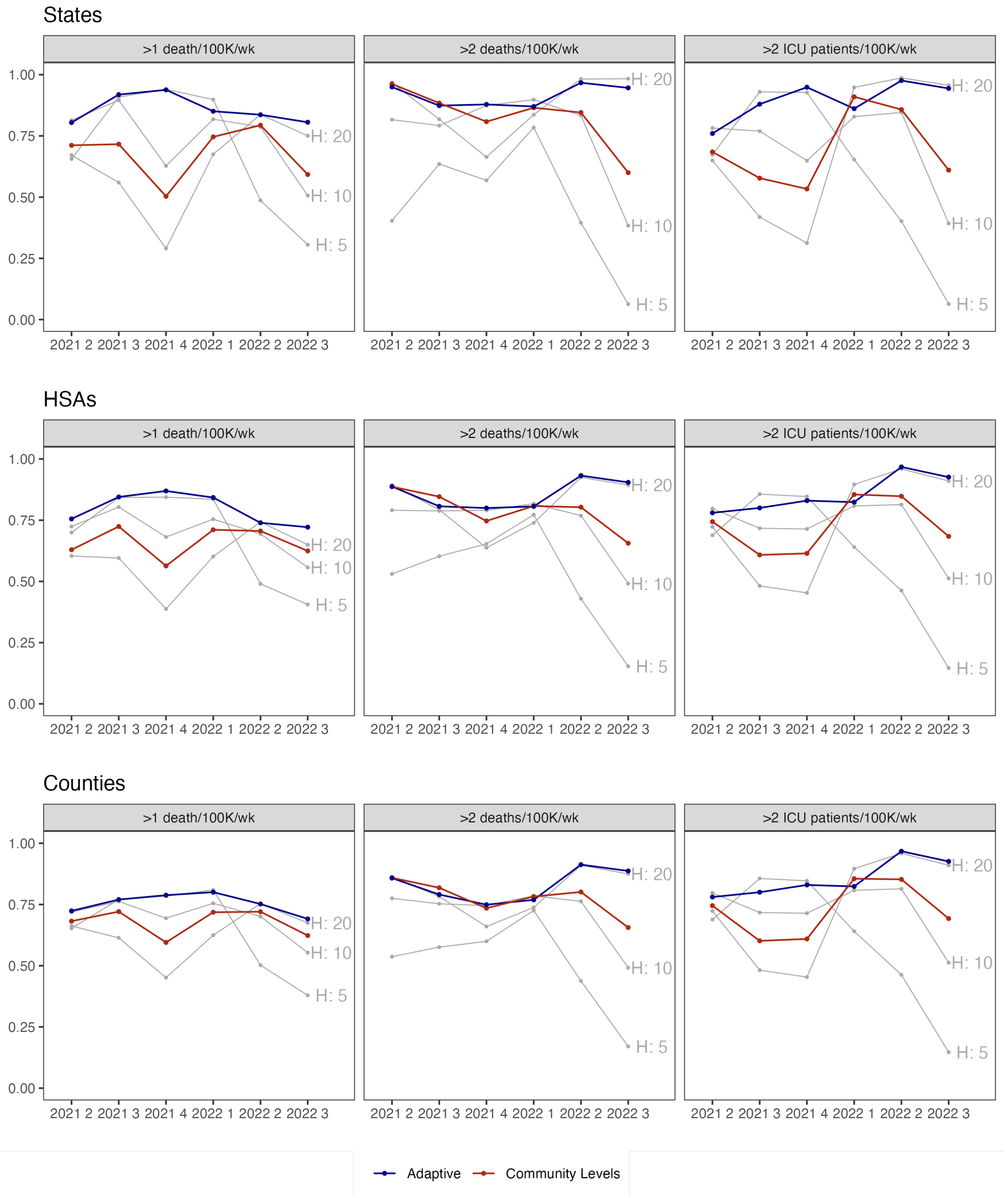


**Fig. S8.** Weighted accuracy by metric. Columns indicate different outcomes. The x-axis indicates quarter, and the y-axis predictive accuracy. Grey lines depict metrics based on new hospital admissions exceeding the row threshold. The red line indicates CDC Community Level and the blue line an adaptive metric (HZ).

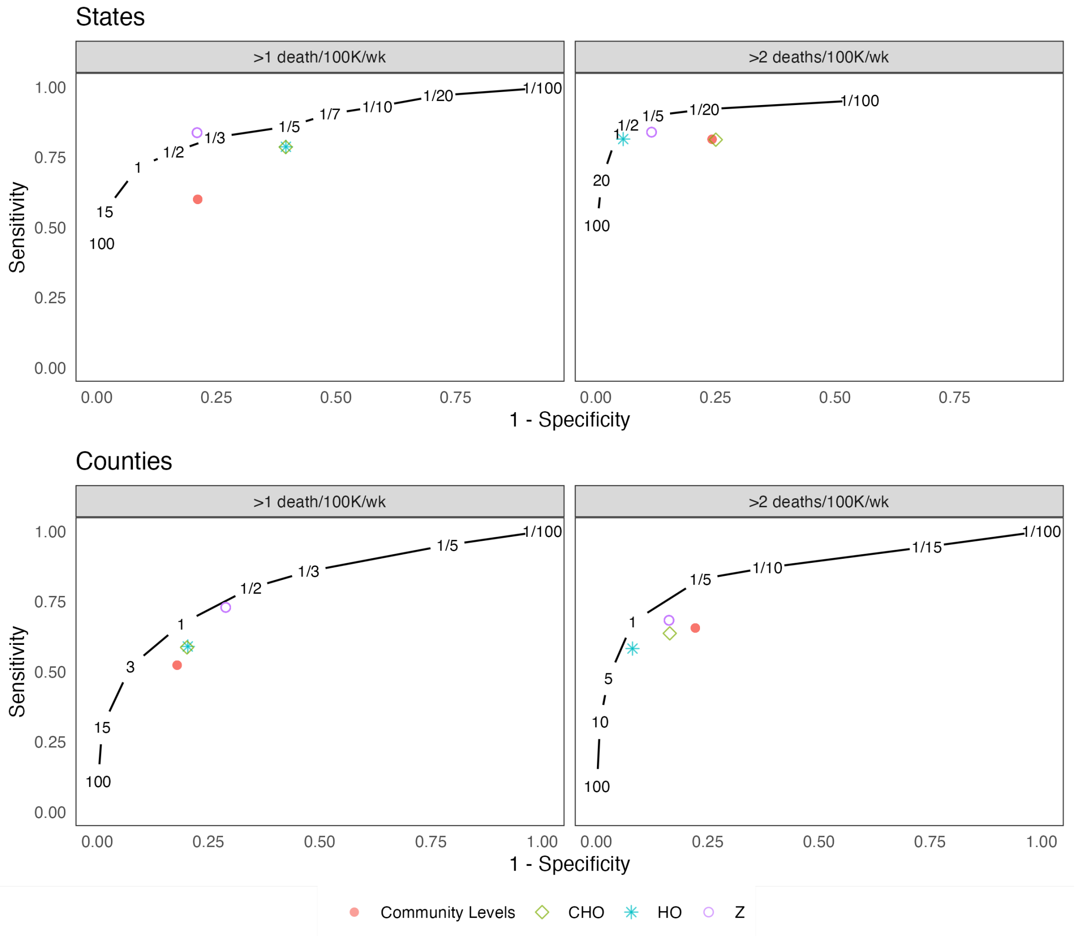


**Fig. S9.** Receiver operating characteristic (ROC) curves for the test period from January 1, 2022 to September 30, 2022. The black line indicates performance of the adaptive metric (HZ) across different values of $wt$, indicating the relative preference for false negatives over false positives. The top plot indicates states and the bottom plot counties.

### States

>1 death/100K/wk

| | Neutral: Training | Neutral: Training MR | Neutral: Test | Neutral: Test MR | Don't cry wolf (0.5x FN): Training | Don't cry wolf (0.5x FN): Training MR | Don't cry wolf (0.5x FN): Test | Don't cry wolf (0.5x FN): Test MR | Better safe than sorry (0.5x FP): Training | Better safe than sorry (0.5x FP): Training MR | Better safe than sorry (0.5x FP): Test | Better safe than sorry (0.5x FP): Test MR |
|---|---|---|---|---|---|---|---|---|---|---|---|---|
| Adaptive: CHOZ | 97 | 4 | 80 | 5 | 97 | 5 | 85 | 2 | 98 | 2 | 76 | 10 |
| Adaptive: CHO | 97 | 4 | 77 | 10 | 97 | 4 | 84 | 2 | 98 | 2 | 70 | 14 |
| Adaptive: HZ | 98 | 3 | 80 | 5 | 98 | 4 | 85 | 1 | 99 | 2 | 77 | 8 |
| Simplified adaptive: HZ | 98 | 4 | 79 | 8 | 98 | 3 | 82 | 5 | 99 | 2 | 78 | 4 |
| Community Levels | 92 | 20 | 66 | 24 | 95 | 12 | 68 | 29 | 90 | 26 | 64 | 24 |
| Z | 93 | 14 | 78 | 8 | 94 | 11 | 76 | 12 | 91 | 16 | 80 | 3 |
| CHO | 98 | 4 | 62 | 41 | 98 | 2 | 57 | 58 | 98 | 4 | 68 | 22 |
| HO | 98 | 4 | 62 | 41 | 98 | 2 | 57 | 58 | 98 | 4 | 68 | 22 |
| CH | 99 | 0 | 49 | 46 | 99 | 0 | 37 | 67 | 99 | 0 | 60 | 23 |
| H | 98 | 3 | 48 | 52 | 98 | 4 | 33 | 77 | 99 | 1 | 63 | 26 |
| C | 99 | 0 | 44 | 46 | 99 | 0 | 31 | 67 | 99 | 0 | 58 | 24 |
| Prevalence | 98 | 3 | 31 | 67 | 98 | 2 | 31 | 73 | 98 | 3 | 31 | 65 |

>2 deaths/100K/wk

| | Neutral: Training | Neutral: Training MR | Neutral: Test | Neutral: Test MR | Don't cry wolf (0.5x FN): Training | Don't cry wolf (0.5x FN): Training MR | Don't cry wolf (0.5x FN): Test | Don't cry wolf (0.5x FN): Test MR | Better safe than sorry (0.5x FP): Training | Better safe than sorry (0.5x FP): Training MR | Better safe than sorry (0.5x FP): Test | Better safe than sorry (0.5x FP): Test MR |
|---|---|---|---|---|---|---|---|---|---|---|---|---|
| Adaptive: CHOZ | 92 | 3 | 91 | 3 | 91 | 8 | 93 | 3 | 94 | 1 | 90 | 5 |
| Adaptive: CHO | 92 | 3 | 93 | 3 | 90 | 9 | 94 | 2 | 94 | 1 | 92 | 3 |
| Adaptive: HZ | 93 | 0 | 92 | 3 | 93 | 0 | 93 | 4 | 94 | 1 | 92 | 3 |
| Simplified adaptive: HZ | 92 | 2 | 94 | 0 | 92 | 2 | 94 | 2 | 93 | 2 | 93 | 1 |
| Community Levels | 92 | 5 | 73 | 36 | 91 | 9 | 68 | 51 | 93 | 4 | 79 | 24 |
| Z | 78 | 18 | 88 | 19 | 82 | 13 | 86 | 24 | 73 | 25 | 91 | 15 |
| CHO | 93 | 1 | 71 | 49 | 94 | 2 | 91 | 6 | 94 | 0 | 78 | 33 |
| HO | 93 | 2 | 91 | 5 | 93 | 0 | 91 | 10 | 94 | 0 | 78 | 35 |
| CH | 93 | 2 | 90 | 6 | 94 | 2 | 91 | 7 | 94 | 1 | 75 | 35 |
| H | 93 | 2 | 90 | 6 | 93 | 0 | 90 | 10 | 93 | 2 | 76 | 38 |
| C | 92 | 6 | 65 | 39 | 91 | 10 | 74 | 27 | 94 | 1 | 63 | 42 |
| Prevalence | 88 | 17 | 9 | 94 | 88 | 19 | 9 | 95 | 88 | 17 | 9 | 92 |

### HSAs

>1 death/100K/wk

| | Neutral: Training | Neutral: Training MR | Neutral: Test | Neutral: Test MR | Don't cry wolf (0.5x FN): Training | Don't cry wolf (0.5x FN): Training MR | Don't cry wolf (0.5x FN): Test | Don't cry wolf (0.5x FN): Test MR | Better safe than sorry (0.5x FP): Training | Better safe than sorry (0.5x FP): Training MR | Better safe than sorry (0.5x FP): Test | Better safe than sorry (0.5x FP): Test MR |
|---|---|---|---|---|---|---|---|---|---|---|---|---|
| Adaptive: CHOZ | 95 | 2 | 73 | 1 | 93 | 2 | 78 | 2 | 96 | 1 | 75 | 0 |
| Adaptive: CHO | 95 | 2 | 69 | 6 | 94 | 1 | 77 | 2 | 96 | 1 | 65 | 11 |
| Adaptive: HZ | 95 | 0 | 73 | 0 | 94 | 1 | 78 | 2 | 96 | 0 | 75 | 0 |
| Simplified adaptive: HZ | 94 | 2 | 72 | 1 | 93 | 3 | 77 | 3 | 96 | 1 | 74 | 1 |
| Community Levels | 89 | 15 | 63 | 13 | 92 | 6 | 68 | 13 | 86 | 26 | 57 | 27 |
| Z | 89 | 9 | 72 | 2 | 91 | 6 | 70 | 12 | 87 | 14 | 74 | 3 |
| CHO | 92 | 7 | 65 | 13 | 94 | 1 | 69 | 18 | 91 | 15 | 61 | 16 |
| HO | 92 | 7 | 65 | 13 | 94 | 1 | 69 | 18 | 91 | 15 | 61 | 16 |
| CH | 94 | 2 | 50 | 31 | 94 | 0 | 44 | 47 | 95 | 3 | 60 | 15 |
| H | 94 | 2 | 51 | 31 | 93 | 1 | 40 | 55 | 95 | 3 | 61 | 15 |
| C | 95 | 2 | 47 | 32 | 94 | 1 | 37 | 54 | 96 | 2 | 58 | 23 |
| Prevalence | 94 | 3 | 39 | 47 | 94 | 1 | 39 | 56 | 94 | 6 | 39 | 47 |

>2 deaths/100K/wk

| | Neutral: Training | Neutral: Training MR | Neutral: Test | Neutral: Test MR | Don't cry wolf (0.5x FN): Training | Don't cry wolf (0.5x FN): Training MR | Don't cry wolf (0.5x FN): Test | Don't cry wolf (0.5x FN): Test MR | Better safe than sorry (0.5x FP): Training | Better safe than sorry (0.5x FP): Training MR | Better safe than sorry (0.5x FP): Test | Better safe than sorry (0.5x FP): Test MR |
|---|---|---|---|---|---|---|---|---|---|---|---|---|
| Adaptive: CHOZ | 88 | 2 | 88 | 0 | 86 | 2 | 91 | 1 | 91 | 0 | 87 | 0 |
| Adaptive: CHO | 87 | 3 | 88 | 0 | 85 | 3 | 91 | 1 | 90 | 2 | 86 | 2 |
| Adaptive: HZ | 88 | 0 | 88 | 1 | 86 | 1 | 90 | 4 | 90 | 1 | 86 | 1 |
| Simplified adaptive: HZ | 85 | 8 | 87 | 2 | 85 | 5 | 91 | 1 | 87 | 10 | 85 | 2 |
| Community Levels | 87 | 4 | 73 | 24 | 86 | 3 | 70 | 37 | 88 | 6 | 75 | 14 |
| Z | 77 | 10 | 83 | 11 | 81 | 5 | 81 | 17 | 73 | 17 | 85 | 9 |
| CHO | 87 | 3 | 78 | 21 | 85 | 0 | 86 | 10 | 89 | 3 | 77 | 17 |
| HO | 87 | 3 | 78 | 21 | 85 | 0 | 86 | 10 | 89 | 3 | 77 | 17 |
| CH | 87 | 2 | 66 | 40 | 85 | 1 | 84 | 15 | 89 | 4 | 58 | 41 |
| H | 87 | 3 | 65 | 41 | 85 | 1 | 84 | 15 | 89 | 3 | 72 | 24 |
| C | 86 | 7 | 49 | 52 | 85 | 5 | 70 | 27 | 90 | 3 | 54 | 42 |
| Prevalence | 83 | 15 | 14 | 86 | 83 | 15 | 14 | 88 | 83 | 17 | 14 | 84 |

### Counties

>1 death/100K/wk

| | Neutral: Training | Neutral: Training MR | Neutral: Test | Neutral: Test MR | Don't cry wolf (0.5x FN): Training | Don't cry wolf (0.5x FN): Training MR | Don't cry wolf (0.5x FN): Test | Don't cry wolf (0.5x FN): Test MR | Better safe than sorry (0.5x FP): Training | Better safe than sorry (0.5x FP): Training MR | Better safe than sorry (0.5x FP): Test | Better safe than sorry (0.5x FP): Test MR |
|---|---|---|---|---|---|---|---|---|---|---|---|---|
| Adaptive: CHOZ | 90 | 0 | 72 | 0 | 89 | 1 | 78 | 1 | 93 | 0 | 71 | 0 |
| Adaptive: CHO | 90 | 2 | 70 | 3 | 89 | 0 | 78 | 2 | 93 | 0 | 65 | 7 |
| Adaptive: HZ | 90 | 0 | 71 | 1 | 89 | 1 | 78 | 2 | 93 | 0 | 71 | 0 |
| Simplified adaptive: HZ | 90 | 3 | 70 | 4 | 88 | 3 | 78 | 3 | 93 | 0 | 71 | 1 |
| Community Levels | 86 | 10 | 64 | 9 | 87 | 3 | 69 | 15 | 85 | 19 | 60 | 21 |
| Z | 83 | 7 | 69 | 4 | 86 | 3 | 67 | 12 | 81 | 13 | 71 | 6 |
| CHO | 89 | 4 | 66 | 9 | 89 | 1 | 69 | 19 | 89 | 9 | 63 | 12 |
| HO | 89 | 4 | 66 | 9 | 89 | 1 | 69 | 19 | 89 | 9 | 63 | 12 |
| CH | 90 | 2 | 50 | 31 | 88 | 1 | 63 | 29 | 92 | 2 | 61 | 12 |
| H | 90 | 2 | 50 | 31 | 88 | 1 | 63 | 29 | 92 | 1 | 61 | 12 |
| C | 90 | 1 | 47 | 33 | 88 | 3 | 36 | 53 | 93 | 0 | 57 | 20 |
| Prevalence | 89 | 3 | 36 | 51 | 89 | 2 | 36 | 58 | 89 | 7 | 36 | 48 |

>2 deaths/100K/wk

| | Neutral: Training | Neutral: Training MR | Neutral: Test | Neutral: Test MR | Don't cry wolf (0.5x FN): Training | Don't cry wolf (0.5x FN): Training MR | Don't cry wolf (0.5x FN): Test | Don't cry wolf (0.5x FN): Test MR | Better safe than sorry (0.5x FP): Training | Better safe than sorry (0.5x FP): Training MR | Better safe than sorry (0.5x FP): Test | Better safe than sorry (0.5x FP): Test MR |
|---|---|---|---|---|---|---|---|---|---|---|---|---|
| Adaptive: CHOZ | 82 | 0 | 86 | 0 | 80 | 1 | 90 | 1 | 86 | 0 | 84 | 0 |
| Adaptive: CHO | 81 | 1 | 86 | 1 | 79 | 3 | 90 | 0 | 86 | 0 | 83 | 2 |
| Adaptive: HZ | 81 | 1 | 86 | 2 | 80 | 2 | 89 | 2 | 86 | 0 | 84 | 1 |
| Simplified adaptive: HZ | 79 | 5 | 86 | 1 | 79 | 3 | 90 | 0 | 83 | 8 | 83 | 1 |
| Community Levels | 82 | 0 | 73 | 23 | 78 | 4 | 71 | 34 | 85 | 2 | 75 | 13 |
| Z | 71 | 7 | 81 | 11 | 75 | 5 | 79 | 16 | 68 | 14 | 83 | 10 |
| CHO | 81 | 0 | 77 | 21 | 80 | 1 | 87 | 6 | 85 | 0 | 76 | 16 |
| HO | 81 | 0 | 77 | 21 | 80 | 0 | 86 | 9 | 85 | 0 | 76 | 16 |
| CH | 81 | 0 | 66 | 38 | 79 | 2 | 86 | 8 | 85 | 0 | 71 | 23 |
| H | 81 | 1 | 65 | 40 | 79 | 2 | 83 | 13 | 85 | 0 | 71 | 23 |
| C | 80 | 4 | 62 | 32 | 77 | 6 | 78 | 15 | 85 | 2 | 55 | 38 |
| Prevalence | 76 | 15 | 15 | 82 | 76 | 18 | 15 | 85 | 76 | 18 | 15 | 79 |

**Fig. S10.** Head-to-head comparison results (omicron training set). The top plots display results from state-level analyses and the bottom plots display results from county-level analyses, both weighted for population. Metrics are displayed on the left, with training data from December 15, 2021-February 15, 2022 and test data from February 16-September 30, 2022. Cells report weighted accuracy and maximum regret (MR) over training and test periods. Rows vary outcomes, and columns vary preferences for false positive versus false negatives, with "neutral" corresponding to unweighted accuracy. Prevalence indicates the proportion of high location-weeks in a given time period.

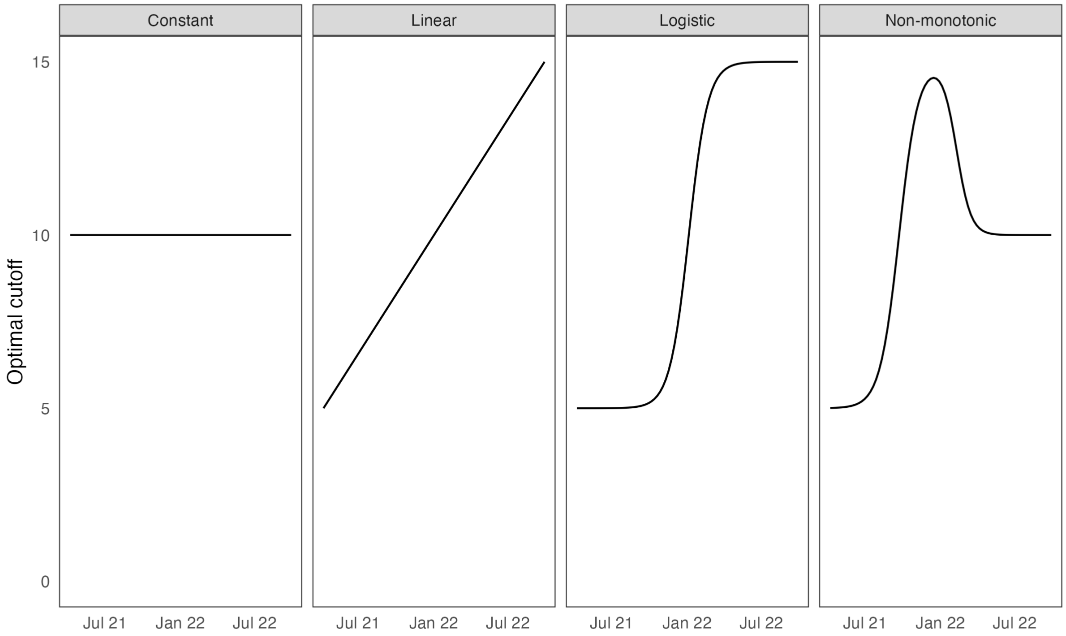


**Fig. S11.** Simulation scenarios. We vary the optimal cutoff for hospitalization to classify a location-week as "high" $(-\beta_0/\beta_1)$ over time in different scenarios. The constant scenario assumes a static relationship between indicators and outcomes, while other scenarios assume a changing relationship.

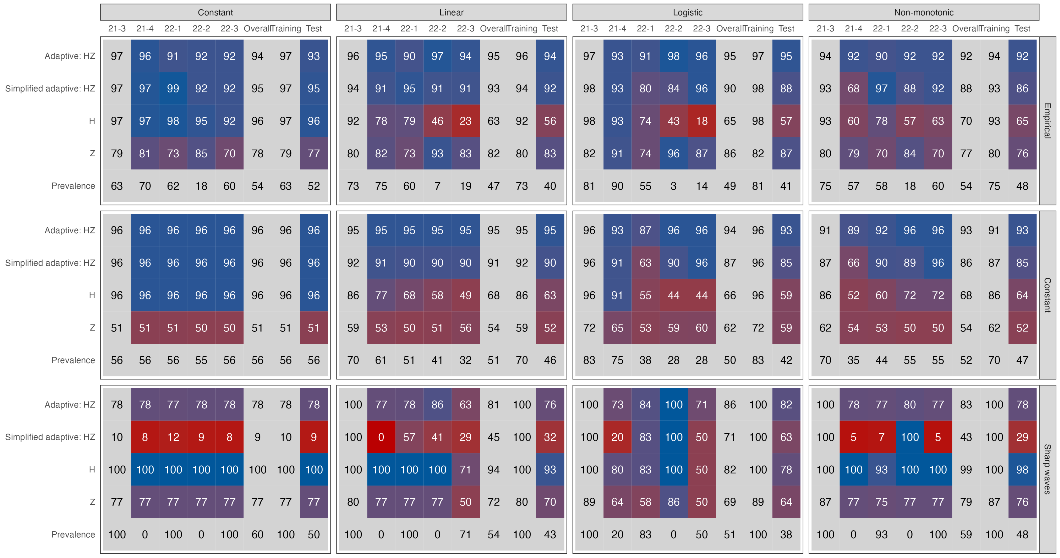


**Fig. S12.** Simulation results. Columns vary the relationship between the input indicator and outcome over time (Figure S11) and rows vary indicator (input) prevalence. Metrics are varied over the y-axis, and 3-week-ahead predictive accuracy is displayed in cells.
